# The estimated health and economic impacts of the introduction of taxation and warning labelling on foods high in fat, sugar or salt in England: A microsimulation study

**DOI:** 10.1101/2025.09.23.25336465

**Authors:** I Gusti Ngurah Edi Putra, Brendan Collin, Zoé Colombet, Rebecca Evans, Ben Amies-Cull, Prof Eric Robinson, Martin O’Flaherty, Chris Kypridemos

## Abstract

**Background:** Foods high in fat, sugar, or salt (HFSS) increase risk of obesity and diet-related illnesses. Some policies, limiting HFSS placements, advertisement, and promotion, have been introduced in the UK to lower HFSS exposure, but evidence remains limited on other policy options and their potential impacts. In this study, we estimated the health and economic impacts of implementing taxation and nutrient warning (NW) labelling on HFSS products.

**Methods:** We used the IMPACT_NCD_, a validated open-cohort microsimulation model, to simulate the health and economic impacts of implementing, for HFSS products, i) 8% taxation, ii) NW labelling, and iii) both combined among English adults aged 30-99 years over 15 years (2026-2040). The IMPACT_NCD_ model utilises nationally representative surveys and routine administrative data to simulate policy and counterfactual scenarios over individuals’ life course within a competing risk framework. Based on available evidence, we estimated the policy impacts based primarily on assumed consumer-driven changes in energy and salt intake, but also examined likely reformulation in sensitivity analyses.

**Findings:** Compared with the current counterfactual scenario (i.e., no taxation, voluntarily implemented multiple traffic light labelling), an 8% tax rate on HFSS products would reduce obesity prevalence by 2.19 percentage points [95% uncertainty intervals (UI): (1.69, 2.73)], prevent 100 000 [95% UI: (65 000, 140 000)] type 2 diabetes mellitus (T2DM), 83 000 [95% UI: (55 000, 130 000)] cardiovascular disease (CVD), 64 000 [95% UI: (48 000, 96 000)] multimorbidity or multiple long-term condition (MLTC) (i.e., Cambridge Multimorbidity Score >1.5) cases, postpone 23 000 [95% UI: (14 000, 37 000)] deaths, and save a total of £53 billion [95% UI: (41, 67)] from a societal perspective (a sum of healthcare, social care, informal care, and productivity) over 15 years. NW labelling was estimated to reduce obesity prevalence by 0.48 percentage points [95% UI: (0.20, 0.85)], prevent 26 000 [95% UI: (13 000, 52 000)] T2DM, 24 000 [95% UIs: (10 000, 41 000)] CVD, 17 000 [95% UI: (8200, 29 000)] MLTC cases, and postpone 6200 [95% UI: (2200, 12 000)] deaths, and save £14 billion [95% UIs: (6.2, 24)]. Both policies implemented together would provide greater impacts with a 2.66 percentage-point [95% UI: (2.09, 3.28)] reduction in obesity prevalence, 130 000 [95% UI: (80 000, 170 000)] T2DM, 110 000 [95% UI: (68 000, 150 000)] CVD, 81 000 [95% UI: (58 000, 110 000)] MLTC cases prevented, and 29 000 [95% UI: (18 000, 45 000)] deaths postponed, and £66 billion [95% UI: (52, 82)] saved. All modelled policies were estimated to be cost-effective and have greater health benefits for individuals in deprived groups. Sensitivity analysis involving both consumer behaviour and industry reformulation to reduce the number of products classed as HFSS yielded similar estimates.

**Interpretation:** This is the first modelled evidence on the health and economic impacts of HFSS taxation and NW labelling. Implementing both taxation and NW labelling as additional measures to reduce HFSS consumption in the UK may provide substantial benefits in addressing obesity and diet-related diseases.

**Funding:** National Institute for Health and Care Research

**Research in context:** *Evidence before this study:* We searched modelling studies on taxation or nutrient warning (NW) labelling on products high in fat, sugar or salt (HFSS) on PubMed from 1^st^ January 2000 to 30^th^ June 2025 using the following search terms: (“food polic*“OR “fiscal polic*” OR tax* OR“label polic*” OR “nutr* label*” OR “food* label*” OR “front of pack label*” OR “front-of-pack label*” OR “warning label*”) AND (*simulation OR “model* study”) based on titles and abstracts, restricted to human subjects. We identified 241 articles. To date, no modelling studies have estimated multiple policy options, such as taxation and NW labelling on all HFSS products in the UK. A handful of studies have reported the substantial impact of taxation on sugary drinks, including in the UK, but limited studies have estimated the impact of taxation on broader unhealthy or HFSS products. One study modelled an 8% tax rate on junk food in New Zealand and reported the policy would reduce the incidence of diabetes mellitus by 7%, coronary heart disease by 3%, and the all-cause mortality rate by 1% over 30 years. Evidence from Mexico suggests greater benefit from doubling the tax rate compared to the currently implemented 8% tax rate on non-essential energy-dense foods. For NW labelling, a previous study estimated that NW labelling on all liable packaged products would reduce obesity prevalence by 2.64 percentage points and prevent 46 000 obesity-related deaths in England over 20 years, but the specific impacts of NW labelling on HFSS products, as well as the broader benefits on different health outcomes and associated economic impacts, have not yet been estimated. A study from Mexico reported that NW labelling could reduce 1.3 million obesity cases or by 4.98 percentage points in its prevalence 5 years post-implementation.

*Added value of this study:* The present study is the first modelled evidence of the health and economic impacts of taxation (an 8% tax rate, informed by the currently implemented tax rate in Mexico) and NW labelling in England. We used the IMPACT_NCD_ – a validated dynamic, stochastic, discrete-time, open-cohort microsimulation model with a competing risk framework at its core – to simulate the health and economic impacts of implementing, for HFSS products, i) taxation, ii) NW labelling, and iii) both combined among English adults aged from 30 to 99 years over a 15-year simulation horizon (2026-2040). We estimated the impacts on a range of health outcomes, including obesity, type 2 diabetes mellitus, cardiovascular diseases, multimorbidity or multiple long-term conditions (MLTCs), and all-cause mortality. Economic impacts were projected from healthcare (medical) and societal (medical, social care, informal care, and productivity) perspectives. We estimated that implementing both policies together would have greater impacts in addressing obesity and diet-related diseases, while also resulting in considerable economic savings. The modelled policies may reduce absolute and relative social health inequalities, as greater benefits were estimated in individuals from deprived groups.

*Implications of all the available evidence:* The present study provides evidence to support the UK Government in considering both HFSS taxation and NW labelling as additional measures to existing policies (restrictions of HFSS placements, advertisements, and promotions) to address obesity and diet-related diseases.

## Introduction

Obesity is a major public health concern in the UK and costs the National Health Service (NHS) over £11.4 billion annually.^1^ The UK Government has measures in place to tackle obesity by targeting food products high in fat, sugar or salt (HFSS). The regulation to restrict HFSS products in key locations (e.g., store entrances) for retail stores (and equivalent key locations online) came into effect on 1 October 2022.^2^ This is followed by restrictions on volume pricing (e.g., multibuy offers) coming into force on 1 October 2025, and related advertising on TV and online on 5 January 2026.^2,3^ Fiscal measures are also plausible, but the current implementation is limited to sugar-sweetened beverages (SSBs) (the Soft Drinks Industry Levy - SDIL) since April 2018.^4^ A controlled interrupted time series study found that SDIL decreased the daily intake of added or free sugar from drinks by 5.3 grams in adults.^5^ A modelling study suggested that SDIL may reduce the prevalence of overweight and obesity by 0.18 to 0.20 percentage points and prevent 12,000 type 2 diabetes mellitus (T2DM) cases and 3,800 cardiovascular disease (CVD) cases over the first 10 years of implementation in the UK, with an estimated healthcare cost saving of £174 million.^4^

In the UK, more than 40% of daily energy intake (700 to 800 kcal) is from HFSS products,^6,7^ while SSBs contribute to 10% of daily free sugar intake (∼5 grams) in adults, which equates to less than 20 kcal per day.^8^ Therefore, greater public health benefits may be achieved if the currently implemented taxation on SSBs (SDIL) is also extended to HFSS products in the UK. A consumer-demand model estimating the price elasticity of HFSS products in Scotland suggested that a 10% tax on HFSS food groups may reduce purchases by 6-10%.^9^ However, evidence on the long-term health and economic impacts of HFSS taxation within the UK context remains limited, despite its importance for informing current public health policies and practices.

Some countries have implemented taxation on unhealthy food. An 8% tax rate was imposed on unhealthy energy-dense food in Mexico in 2014, and this taxation led to a reduction in sales of taxed food from 4.8% in the first year to 7.4% in the second year after the implementation.^10^ In 2011, Hungary introduced “a public health product tax”, an excise tax on products exceeding the threshold of sugar or salt content, with a relative tax rate ranging from 5% to 10% for most products.^11^ Based on experience in Hungary, taxation effectively reduced the consumption of taxed food in the short term, but a complex intervention is required to attain sustainable healthy diets.^11^ Therefore, an additional intervention aimed at encouraging individuals to make informed and healthier choices, such as HFSS nutrient warning (NW) labelling, alongside taxation, could be considered as complementary measures to reduce HFSS consumption.

The UK Government launched a consultation on front-of-pack nutrition labelling in 2020 and considered Chile’s NW labelling (Appendix Figure 1A) as an alternative to the current implemented voluntary multiple traffic light (MTL) labelling (Appendix Figure 1B).^12^ Implementing NW labelling on HFSS products may have greater impacts than MTL labelling in encouraging consumers to select healthier options.^13^ This aligns with findings from a network meta-analysis,^14^ suggesting that the effect of NW outperformed MTL labelling in reducing total energy purchased by 6.4% (95% CI: [0.4; 12.5]). A recent modelling study estimated greater impacts of implementing mandatory NW compared to MTL labelling on liable packaged products in preventing obesity and obesity-related mortality in England without widening current health inequalities.^15^

There is a dearth of evidence on the likely population health impacts of taxation and NW labelling on HFSS products. The present study aimed to estimate the population-level impacts of taxation and NW labelling on HFSS products, both separately and in combination, on obesity (body mass index – BMI ≥ 30 kg/m^2^), T2DM, CVDs, multimorbidity or multiple long-term conditions (MLTCs), all-cause mortality, and related economic impact in England, as well as the socioeconomic (SES)-related health inequalities in the estimated impacts.

## Methods

We used the IMPACT_NCD_ model (see the model description^16^ and appendix p 3) to estimate the impacts of taxation and NW labelling on HFSS products over a 15-year modelling horizon (2026-2040) in England. IMPACT_NCD_ is a validated dynamic, stochastic, discrete-time, open-cohort microsimulation model that has been used to estimate the health and economic impacts of public health policies on non-communicable diseases (NCDs) in different countries.^17,18^ The model employs a competing risk framework, enabling individuals to die from other modelled diseases when a specific policy may extend their lifespan by reducing the risk of death from the disease of interest.

We simulated the impact of HFSS taxation and NW labelling in the population of England aged 30 to 99 years, as we estimated the policy impacts on a range of diseases (e.g., T2DM, CVDs, MLTCs) that are rare in younger age groups. Three main scenarios (taxation, NW labelling, and both combined) were modelled. We followed Mexico’s tax rate of 8%.^19^ This tax rate is comparable to a range of 5% to 10% for most unhealthy products in Hungary.^11^ For HFSS NW labelling, our scenario was informed by evidence from a meta-analysis (see below).^14^ In the final scenario, we modelled the impacts of both policies combined, assuming they are independent (see below). The scenarios above were compared to a counterfactual baseline scenario representing the current situation or policy (no taxation on HFSS products and the currently implemented voluntary MTL labelling policy).

For each scenario, we modelled the policy effects on reducing energy (kcal) and salt intake (grams) that would immediately impact BMI and systolic blood pressure (SBP), respectively. Changes in BMI and SBP affected the probability of modelled disease outcomes. We implemented taxation and NW labelling on all HFSS products (defined using the UK nutrient profiling model - NPM),^6^ consumed from non-out-of-home food businesses (e.g., supermarkets, retailers) (appendix p 5).

### The effects of taxation and NW labelling on HFSS products

#### Effect of taxation

We estimated the effect of an 8% tax rate through consumer responses to price changes. We used empirical evidence from Scotland, suggesting that a 10% tax on HFSS food groups may result in a 6-10% reduction in HFSS products purchased (assuming the quantity purchased equates to its consumption).^9^ Therefore, an 8% tax rate equates to a 4.8 to 8% reduction in consumption of HFSS products, applied equally to both energy and salt intake (appendix p 6). This assumed effect aligns with findings from a meta-analysis by Afshin et al.,^20^ suggesting that a 10% price increase reduced consumption of unhealthy foods (excluding SSBs and fast food) by 9% (95% CI: [6, 12]). The assumed effect of 4.8% to 8% for an 8% tax rate is consistent with empirical data from post-taxation in Mexico, which showed reductions in sales of taxed food ranging from 4.8% in the first to 7.4% in the second year post-implementation.^10^ We assumed effects on consumer responses would be similar across SES groups due to limited direct evidence, supported by findings from sugary drink taxation.^21^ However, we conducted a sensitivity analysis assuming differential effects of taxation by income groups (see below). A complete pass-through rate was assumed, following an assumption in a previous modelling study^22^ and evidence of average pass-through rates of 105-108% for SDIL^23^. Based on evidence from the same study used to inform the taxation effect,^9^ we also assumed reductions in non-HFSS products, such as fruit and vegetable consumption by 2% and 5%, respectively, due to a 10% tax rate (equivalent reductions for an 8% tax rate: 1.6% and 4%, respectively) in addition to the taxation effects on energy and salt intake. These reductions in fruit and vegetable consumption would increase the risk of CVDs. We conducted a sensitivity analysis, not including reductions in fruit and vegetables. We did not consider compensation (i.e., reduced intake being partially offset by intake from non-taxed foods), following empirical evidence of no impacts on untaxed food in Mexico.^19^

#### Effect of NW labelling

We used evidence from a network meta-analysis by Song et al.,^14^ suggesting that NW labelling would reduce energy by 3.8% (95% CI: [0.9%, 6.8%]) and salt by 2.1% (95% CI: [-4.0%, 8.2%]) per 100 grams of food purchased (assuming the quantity of food purchased equates to its consumption), but may not outperform MTL labelling. This is consistent with a meta-analysis by Crocker et al.^24^ demonstrating similar effects between NW and MTL labelling (appendix p 8). As 75% of all the packaged products currently feature MTL labelling in the UK,^15,25^ we assumed that implementing NW labelling would only affect the remaining HFSS products without MTL labelling (25%). We assumed similar effects across sociodemographic characteristics, following the current literature.^14,15,26^

#### Combined effects

We assumed that taxation and NW labelling policies would have independent effects; therefore, we modelled the simultaneous effects of the policies using a multiplicative approach. We followed the SimSmoke tobacco control policy simulation model approach^27^ used to estimate the impact of different policies combined (e.g., taxation, health warning) (appendix p 10).

#### Sensitivity analyses

In our primary analysis, we opted to focus on consumer responses only, as there is greater uncertainty when estimating the likely degree of reformulation. In sensitivity analyses, we estimated the policy impacts through both consumer response and reformulation (i.e., reduction in energy and salt content of HFSS products). For taxation, we assumed a 6% reduction in energy content and a 15% reduction in salt content of HFSS products, calculated using evidence from the Institute for Fiscal Studies (appendix p 7).^28^ We used findings from a meta-analysis^29^ on potential reduction in energy and salt contents due to reformulation, by 0.9% (95% CI: [–3.1, 4.9]) and 8.9% (95% CI: [0.6%, 17.3%]), respectively. We assumed reformulation coverages of 40% for taxation and 20% for NW labelling, following evidence from Mexico and Chile, respectively. We adjusted the policy coverages when examining the effects of both consumer response and reformulation (i.e., product reformulation reduces the proportion of liable HFSS products subjected to taxation or NW labelling; appendix pp 7-10).

We conducted additional sensitivity analyses on consumer-driven changes in energy and salt intake. Due to limited evidence within the UK context, we assumed a gradient in tax effects on reducing taxed food purchases by income levels based on evidence from Mexico’s 8% taxation: low (10.2%), middle (5.8%), and high (0%) household income groups (with an overall effect of 5.1%).^19^

Following a previous modelling study,^15^ we conducted a sensitivity analysis based on reductions in total energy and salt intake (as opposed to reduction per 100-gram unit) for NW labelling. Song et al.,^14^ reported that NW labelling reduced total energy (kcal) purchased by 12.9% (95% CI: [8.0%; 17.9%]) and outperformed MTL labelling by 6.4% (95% CI: [0.4%; 12.5%]) in reducing total energy purchased. The effects on salt were not statistically significant for i) NW labelling vs. control (7.8%; 95% CI: [-1.2%; 16.8%]) and ii) NW vs. MTL labelling (1.3%; 95% CI: [-7.7%; 10.3%]) in reducing total salt.

### Data sources

A close-to-reality synthetic population of England was created using data from the Health Survey for England (HSE) (to inform individual traits and trends in risk factors), Office for National Statistics (ONS) (population projection by age, sex, and socioeconomic status), and Clinical Practice Research Datalink Aurum, Hospital Episode Statistics, and ONS mortality records (to inform trends in disease-specific prevalence and mortality) (appendix p 3). Using the National Diet and Nutrition Survey (NDNS) years 1 (2009) to 11 (2019), we fitted generalised additive models for location, shape, and scale (GAMLSS) models to estimate the trends of energy (kcal), salt (grams) intake, and food weight (grams) from HFSS products throughout the simulation horizon (appendix p 5).

### Model engine

The IMPACT_NCD_ simulated trends of non-communicable disease risk factors, including BMI, SBP, smoking status, second-hand smoking, fruit and vegetable consumption, alcohol intake, total cholesterol and active days based on data from HSE from 2003 to 2014 (Appendix Figure 2). GAMLSS were used to model trends in these risk factors conditional on some functions of year, sex, age, ethnicity, and deprivation level as independent variables (see IMPACT_NCD_ documentation^16^, appendix p 3). This enables us to model the impact of taxation and NW labelling on health outcomes through reduced energy and salt intake while accounting for other relevant disease risk factors in a competing risk framework. Under the policy scenarios, we calculated changes in energy and salt intake based on baseline intake and the policy effects. Changes in energy consumption were then transformed into equivalent changes in BMI, guided by a dynamic model of weight change introduced by Hall.^30,31^ We also accounted for the immediate impact of BMI changes on SBP (appendix p 11). To translate changes in salt into SBP, we used a meta-regression equation reported in a meta-analysis of 103 trials by Mozaffarian, Fahimi ^32^ (appendix p 11). The IMPACT_NCD_ assumed multiplicative risk effects to combine the impact of changes in multiple risk factors on the probability of developing a modelled disease. Based on BMI and SBP changes, we estimated subsequent changes in risk of CVDs, as well as other diseases, to inform MLTCs with a mean lag time of 4-5 years between most exposure and disease pairs. The IMPACT_NCD_ used evidence from high-quality meta-analyses to model changes in risk of developing modelled diseases due to changes in risk factors (see IMPACT_NCD_ documentation^16^ and appendix pp 11-12).

### Model outputs and uncertainty

We estimated the health impacts of the policies on cases and case-years (i.e., the number of cases and the length of time the condition) prevented or postponed of obesity, type 2 DM, CVDs (a sum of coronary heart disease (CHD), stroke, atrial fibrillation, heart failure), MLTCs, and all-cause mortality in England. We used the Cambridge Multimorbidity Score (CMS) to define the level of multimorbidity or MLTCs.^33^ In brief, 20 conditions: “dementia, cancer, chronic obstructive pulmonary disease, atrial fibrillation, heart failure, constipation, chronic pain, epilepsy, stroke/transient ischaemic attack, DM (type 1 or 2), alcohol problems, psychosis/bipolar disorder, chronic kidney disease, anxiety/depression, CHD, connective tissue disorders, irritable bowel syndrome, asthma, hearing loss, and hypertension”, were assigned a weight or score based on the impacts of illness on the use of primary care, emergency health services, and risk of death.^16,33^ We defined MLTCs as a CMS score >1.5 (e.g., as in ^34^). We calculated quality-adjusted life years (QALYs) by multiplying the predicted years of life lived by disease-specific utility weights (appendix p 12). Based on the economic evaluation perspective, economic impacts were estimated from healthcare (i.e., medical) and societal (i.e., a sum of medical, social care, informal care, and productivity) perspectives (appendix pp 12-15).^18^ We also calculated net monetary benefit (NMB), cost-benefit ratio (CBR), and incremental cost-effectiveness ratio (ICER). We applied a discount rate of 3.5% per annum from 2026 to QALYs and costs based on guidance from the UK Treasury (appendix pp 12-18, Appendix Tables 1 and 2).

**Table 1.** Modelling scenarios and key assumptions.

| **Main analysis (consumer response)** |  |
| --- | --- |
| Taxation | An 8% tax rate leads to a consumer-driven reduction of 4.8 to 8% in energy and salt intake from HFSS products. |
| Nutrient warning labelling | NW labelling results in a consumer-driven reduction of 3.8% (95% CI: [0.9%, 6.8%]) in energy intake and 2.1% (95% CI: [-4.0%, 8.2%]) in salt intake (per 100 grams of food) for the remaining 25% HFSS products (without MTL). |
| Taxation and nutrient warning labelling | The combined effects of both taxation and NW labelling (as above) in reducing energy and salt intake from HFSS products. |
| **Sensitivity analysis (consumer response and reformulation)** |  |
| Taxation | An 8% tax rate leads to a consumer-driven reduction of 4.8 to 8% in energy and salt intake from HFSS products.<br>Reformulation due to taxation reduces energy and salt content from HFSS products by 6% and 15%, respectively. |
| NW labelling | NW labelling results in a consumer-driven reduction of 3.8% (95% CI: [0.9%, 6.8%]) in energy intake and 2.1% (95% CI: [-4.0%, 8.2%]) in salt intake (per 100 grams of food) for the remaining 25% HFSS products (without MTL).<br>Reformulation due to NW labelling reduces energy and salt content from HFSS products by 0.9% (95% CI: [-3.1, 4.9]) and 8.9% (95% CI: [-17.3%, -0.6%]), respectively. |
| Taxation and nutrient warning labelling | The combined effects of both taxation and NW labelling (as above) in reducing energy and salt intake from HFSS products. |
| **Sensitivity analysis (consumer response)** |  |
| Taxation | Differential taxation effects on consumer-driven reduction in energy and salt intake by income levels: low (10.2%), middle (5.8%), and high (0%) household income groups. |
| Nutrient warning labelling | NW labelling reduces consumer-driven energy intake by 6.4% (95% CI: [0.4%; 12.5%]) in 75% of HFSS products featuring MTL labelling and by 12.9% (95% CI: [8.0%; 17.9%]) in the remaining 25% (without MTL).<br>NW labelling reduces consumer-driven salt intake by 1.3% (95% CI: [-7.7%; 10.3%]) in 75% of HFSS products featuring MTL labelling and by 7.8% (95% CI: [-1.2%; 16.8%]) in the remaining 25% (without MTL). |

**Table 2.**
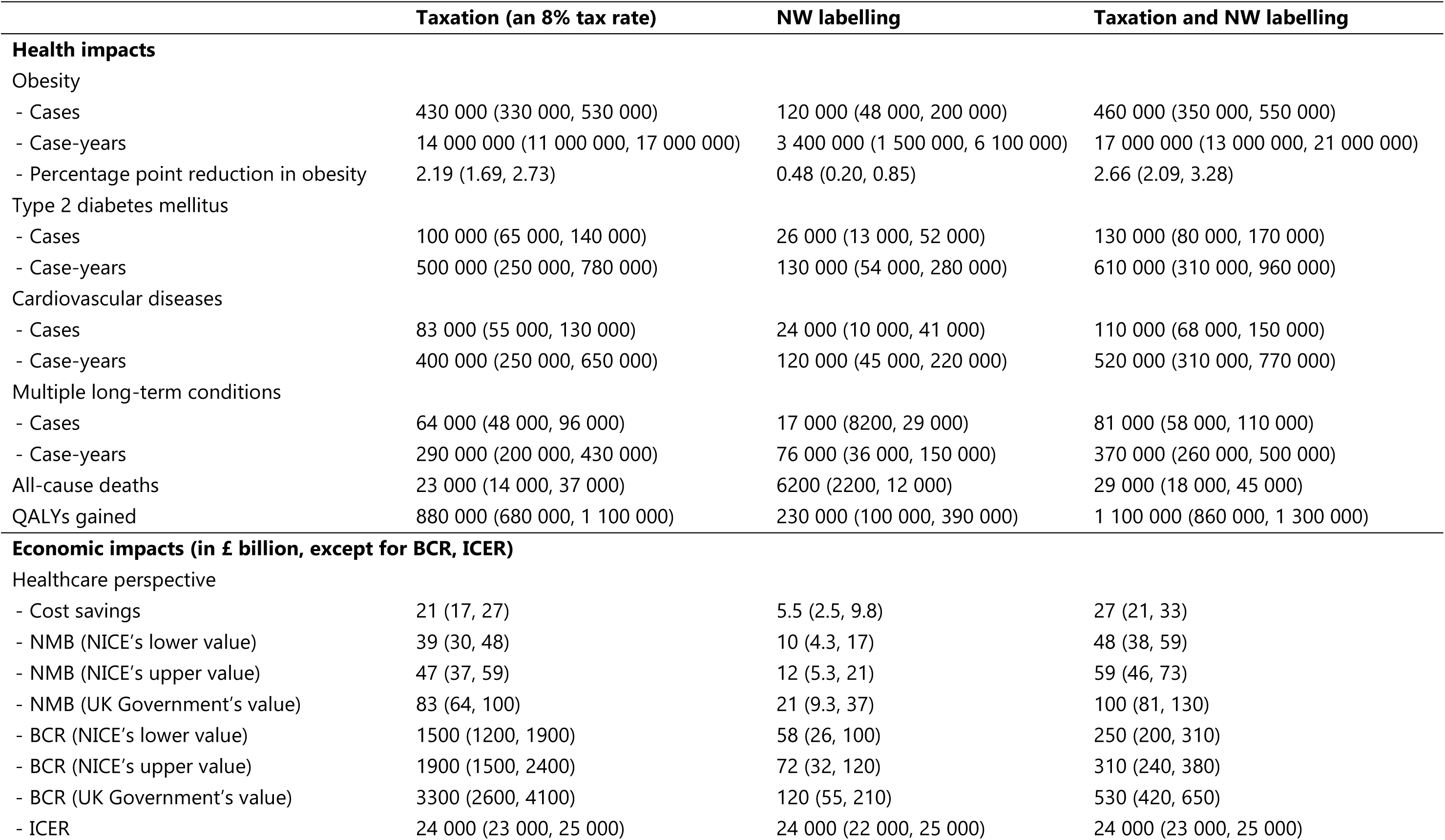
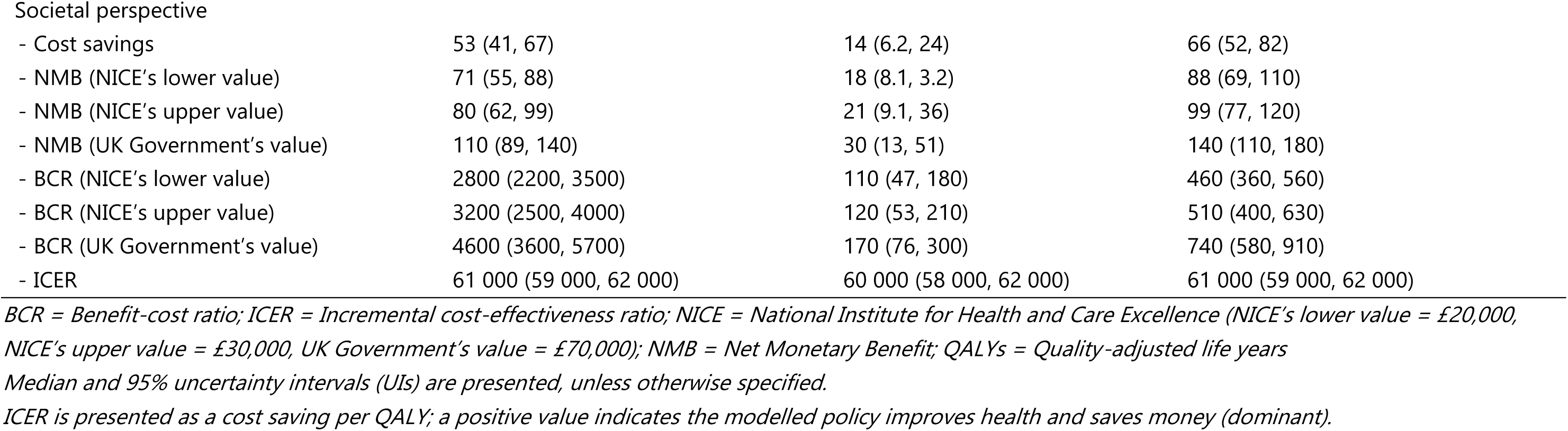
Estimated health and economic impacts of an 8% tax rate, NW labelling, and both policies combined on HFSS food based on consumer responses (2026 – 2040)

|  | **Taxation (an 8% tax rate)** | **NW labelling** | **Taxation and NW labelling** |
| --- | --- | --- | --- |
| **Health impacts** |  |  |  |
| Obesity |  |  |  |
| - Cases | 430 000 (330 000, 530 000) | 120 000 (48 000, 200 000) | 460 000 (350 000, 550 000) |
| - Case-years | 14 000 000 (11 000 000, 17 000 000) | 3 400 000 (1 500 000, 6 100 000) | 17 000 000 (13 000 000, 21 000 000) |
| - Percentage point reduction in obesity | 2.19 (1.69, 2.73) | 0.48 (0.20, 0.85) | 2.66 (2.09, 3.28) |
| Type 2 diabetes mellitus |  |  |  |
| - Cases | 100 000 (65 000, 140 000) | 26 000 (13 000, 52 000) | 130 000 (80 000, 170 000) |
| - Case-years | 500 000 (250 000, 780 000) | 130 000 (54 000, 280 000) | 610 000 (310 000, 960 000) |
| Cardiovascular diseases |  |  |  |
| - Cases | 83 000 (55 000, 130 000) | 24 000 (10 000, 41 000) | 110 000 (68 000, 150 000) |
| - Case-years | 400 000 (250 000, 650 000) | 120 000 (45 000, 220 000) | 520 000 (310 000, 770 000) |
| Multiple long-term conditions |  |  |  |
| - Cases | 64 000 (48 000, 96 000) | 17 000 (8200, 29 000) | 81 000 (58 000, 110 000) |
| - Case-years | 290 000 (200 000, 430 000) | 76 000 (36 000, 150 000) | 370 000 (260 000, 500 000) |
| All-cause deaths | 23 000 (14 000, 37 000) | 6200 (2200, 12 000) | 29 000 (18 000, 45 000) |
| QALYs gained | 880 000 (680 000, 1 100 000) | 230 000 (100 000, 390 000) | 1 100 000 (860 000, 1 300 000) |
| **Economic impacts (in £ billion, except for BCR, ICER)** |  |  |  |
| Healthcare perspective |  |  |  |
| - Cost savings | 21 (17, 27) | 5.5 (2.5, 9.8) | 27 (21, 33) |
| - NMB (NICE's lower value) | 39 (30, 48) | 10 (4.3, 17) | 48 (38, 59) |
| - NMB (NICE's upper value) | 47 (37, 59) | 12 (5.3, 21) | 59 (46, 73) |
| - NMB (UK Government's value) | 83 (64, 100) | 21 (9.3, 37) | 100 (81, 130) |
| - BCR (NICE's lower value) | 1500 (1200, 1900) | 58 (26, 100) | 250 (200, 310) |
| - BCR (NICE's upper value) | 1900 (1500, 2400) | 72 (32, 120) | 310 (240, 380) |
| - BCR (UK Government's value) | 3300 (2600, 4100) | 120 (55, 210) | 530 (420, 650) |
| - ICER | 24 000 (23 000, 25 000) | 24 000 (22 000, 25 000) | 24 000 (23 000, 25 000) |
| Societal perspective |  |  |  |
| - Cost savings | 53 (41, 67) | 14 (6.2, 24) | 66 (52, 82) |
| - NMB (NICE's lower value) | 71 (55, 88) | 18 (8.1, 3.2) | 88 (69, 110) |
| - NMB (NICE's upper value) | 80 (62, 99) | 21 (9.1, 36) | 99 (77, 120) |
| - NMB (UK Government's value) | 110 (89, 140) | 30 (13, 51) | 140 (110, 180) |
| - BCR (NICE's lower value) | 2800 (2200, 3500) | 110 (47, 180) | 460 (360, 560) |
| - BCR (NICE's upper value) | 3200 (2500, 4000) | 120 (53, 210) | 510 (400, 630) |
| - BCR (UK Government's value) | 4600 (3600, 5700) | 170 (76, 300) | 740 (580, 910) |
| - ICER | 61 000 (59 000, 62 000) | 60 000 (58 000, 62 000) | 61 000 (59 000, 62 000) |

*BCR = Benefit-cost ratio; ICER = Incremental cost-effectiveness ratio; NICE = National Institute for Health and Care Excellence (NICE's lower value = £20,000,* *NICE's upper value = £30,000, UK Government's value = £70,000); NMB = Net Monetary Benefit; QALYs = Quality-adjusted life years* *Median and 95% uncertainty intervals (UIs) are presented, unless otherwise specified.* *ICER is presented as a cost saving per QALY; a positive value indicates the modelled policy improves health and saves money (dominant).*

Following a previous study,^35^ we calculated two regression-based metrics, the “absolute equity slope index” and the “relative equity slope index”, using the quintile groups of Index of Multiple Deprivation (QIMD) as an SES measure, taking into account the population distribution by QIMD. We focused on QALYs as this provides a comprehensive view of how the modelled policies reduce burden of a range of diseases within a competing risk framework. The former index indicates the policy impact on absolute inequality (i.e., the number of QALYs gained in most deprived compared to least deprived areas). The latter accounts for pre-existing QIMD gradient of disease burden and represents the policy impact on relative inequality (i.e., positive values indicate the policy tackles relative inequality).

We ran 100 iterations using a Monte Carlo method implemented in the IMPACT_NCD_ to incorporate uncertainties of all the model parameters (appendix p 18 and Appendix Tables 3 and 4). Outputs were reported as medians and 95% uncertainty intervals (UIs) up to two significant figures. UIs indicate a plausible range of estimated impacts after accounting for uncertainties in the underlying data, and these are not equivalent to confidence intervals.

## Results

### The impacts of taxation and NW labelling

Table 2 presents the projected health and economic impacts of an 8% tax rate on HFSS food through the assumed change in consumer response in England. Taxation was estimated to prevent 430 000 obesity cases [95% UI: (330 000, 530 000)] and reduce the prevalence of obesity by 2.19 percentage points [95% UI: (1.69, 2.73)] over the next 15 years (2026 – 2040). The taxation could also prevent 100 000 T2DM cases [95% UI: (65 000, 140 000)], 83 000 CVD cases [95% UI: (55 000, 130 000)], and 64 000 MLTC cases (CMS score >1.5) [95% UI: (48 000, 96 000)]. We projected that 23 000 fewer deaths [95% UI: 14 000 to 37 000] would occur if the taxation were implemented. Through reductions in relevant risk factors (BMI, SBP) and subsequent health conditions, 880 000 QALYs could be gained over 15 years with the 8% tax on HFSS products. We also estimated that £53 billion could be saved over 15 years. A sensitivity analysis excluding reductions in fruits and vegetables showed similar findings (Appendix Table 5). We estimated smaller impacts when assuming differential taxation effects by income levels due to a smaller overall assumed taxation effect (Appendix Table 6).

We estimated that implementing NW labelling over the next 15 years (2026 – 2040) would prevent 120 000 obesity cases [95% UI: (48 000, 200 000)], or a 0.48 percentage point reduction [95% UI: (0.20, 0.85)] (Table 2). The NW labelling was also estimated to prevent 26 000 T2DM [95% UI: (13 000, 52 000)], 24 000 CVD [95% UI: (10 000, 41 000)], and 17 000 MLTC cases [95% UI: (8200, 29 000)]. NW labelling would postpone 6200 deaths [95% UI: 2200 to 12 000]. We estimated 230 000 QALYs could be gained over 15 years by implementing NW labelling on HFSS products. The policy could save £14 billion by 2040. We estimated greater impacts when reductions in total energy and salt intakes were modelled (as opposed to reductions per 100 grams of food) (Appendix Table 7).

Larger impacts were estimated from implementing both taxation and NW labelling (Table 2). We estimated 460 000 [95% UI: (350 000, 550 000)] obesity cases could be prevented, contributing to a reduction in obesity prevalence by 2.66 percentage points [95% UI: (2.09, 3.28)]. In addition, 130 000 T2DM cases [95% UI: (80 000, 170 000)], 110 000 CVD cases [95% UI: (68 000, 150 000)], and 81 000 MLTC cases [95% UI: (58 000, 110 000)] could be prevented if both policies were implemented. The policies combined would result in 29 000 (18 000, 45 000) fewer deaths, 1 100 000 QALYs gained and a total of £66 billion saved, over 15 years.

We are likely to underestimate NMB, BCR, and ICER for i) taxation and ii) combined taxation and NW labelling because we did not include tax revenue due to limited product-level data. Nevertheless, all the modelled policy scenarios were cost-saving and cost-effective because the BCR is > 1 (greater economic benefits than policy costs), and the ICER is dominant (a cost saving per QALY).

### Socioeconomic inequalities in the impacts

Under the assumption of similar effects across SES groups for the main analysis, as supported by previous studies,^14,15,21,26^ Figures 1, 2, and 3 present evidence across the policy scenarios (8% tax rate, NW labelling, and both combined) showing more obesity, type 2 DM, and MLTCs cases prevented and QALYs gained in the most deprived (QIMD 1) compared to the least deprived (QIMD 5), but the UIs overlapped. However, overlapping UIs are not evidence for a lack of statistical significance because the comparisons (QIMD 5 vs. 1) are not independent. The number of CVD cases and deaths postponed generally appeared similar between these two quintiles. We projected greater cost savings in most deprived areas (Figure 4). Our estimates of the absolute and relative equity slope indexes for QALYs gained, accounting for population distribution, suggest that the modelled policies may reduce both absolute and relative inequalities (Appendix Table 8). When assuming differential taxation effects by income groups (as opposed to the same effect in the main analysis), we found stronger evidence on reducing these inequalities (Appendix Table 6).

**Figure 1.**
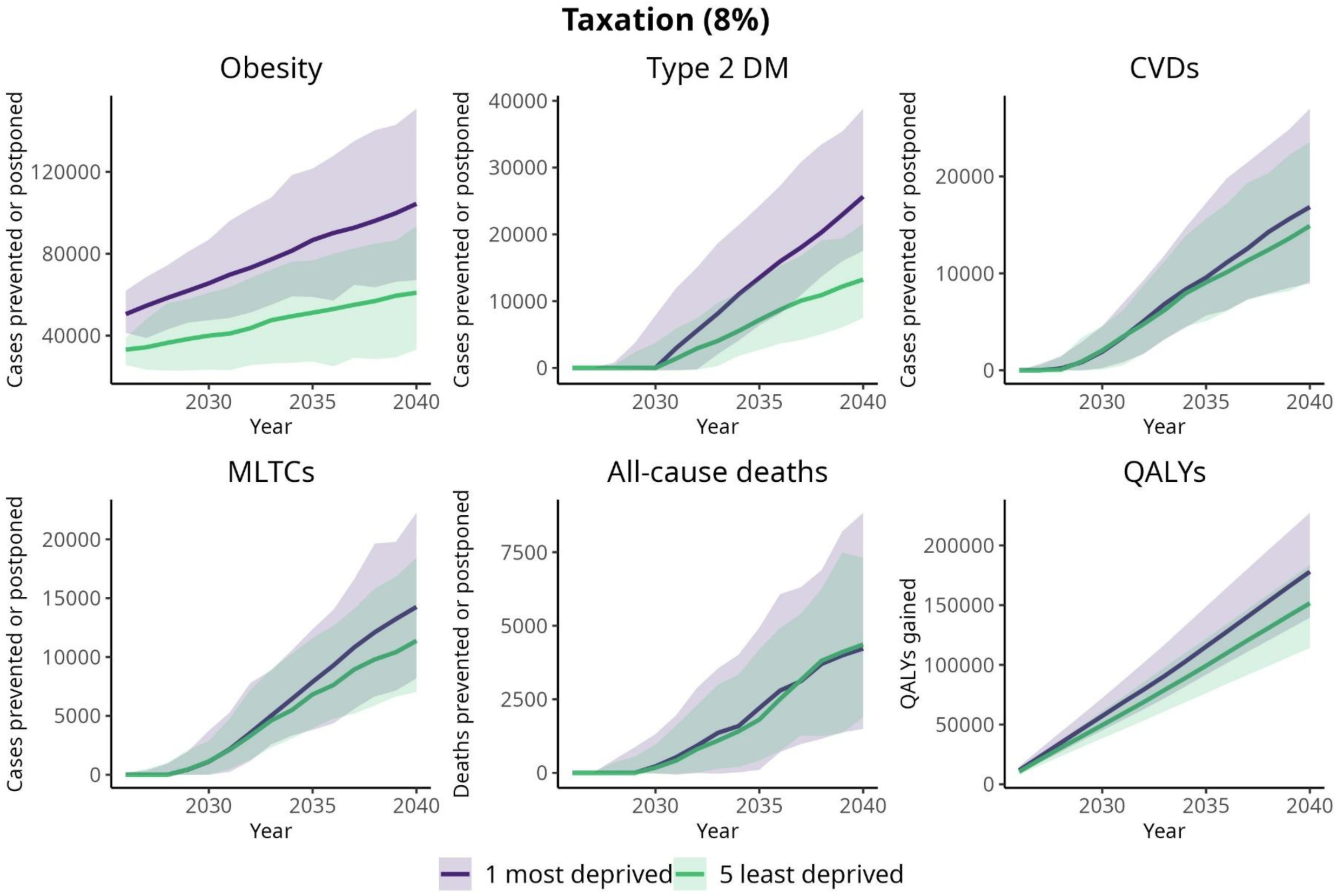
Cumulative cases or deaths prevented or postponed and QALYs gained (median; 95% UI) due to taxation (8%) on HFSS food based on consumer response by QIMD (2026 – 2040)

**Figure 2.**
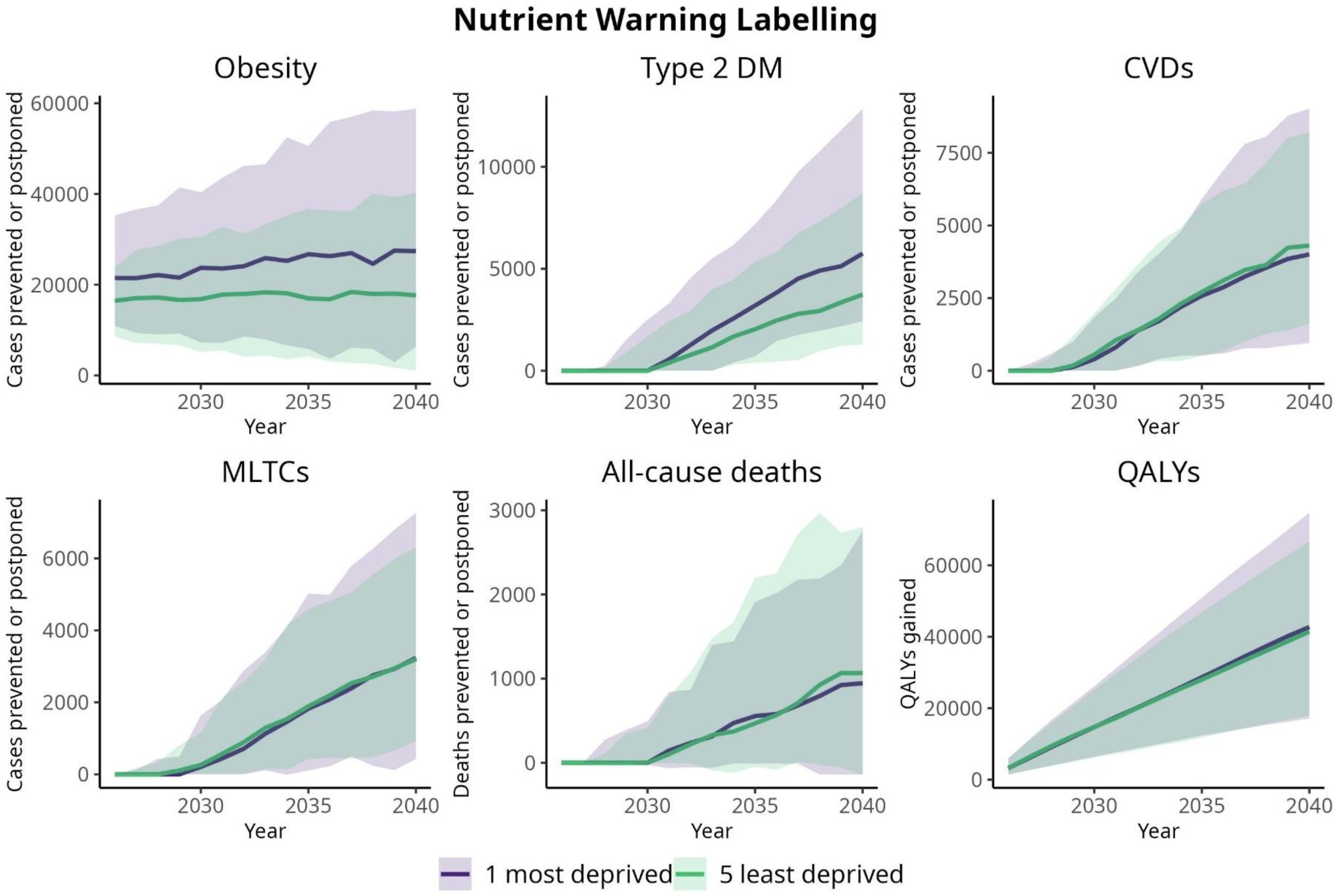
Cumulative cases or deaths prevented or postponed and QALYs gained (median; 95% UI) due to NW labelling on HFSS food based on consumer response by QIMD (2026 – 2040)

**Figure 3.**
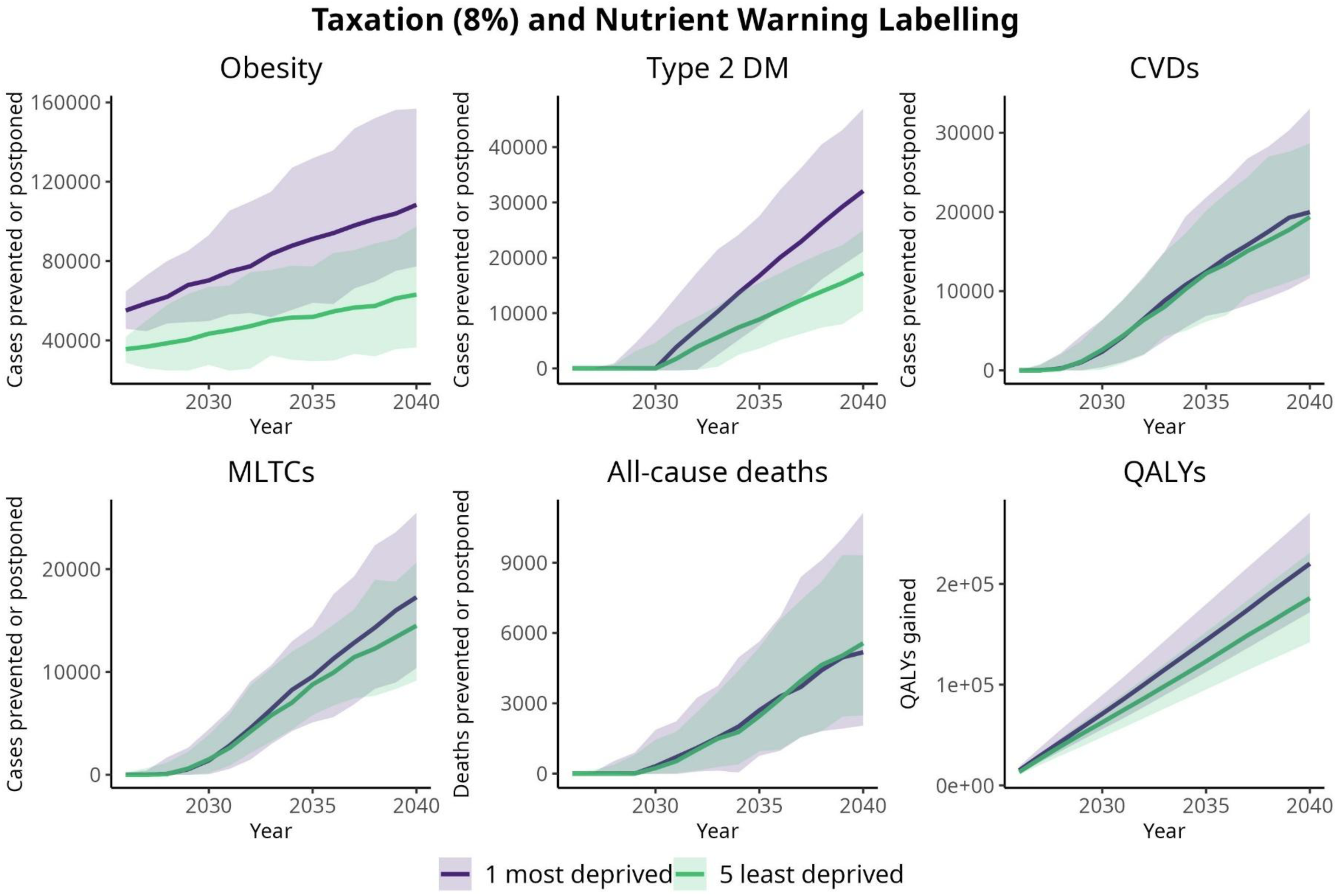
Cumulative cases or deaths prevented or postponed and QALYs gained (median; 95% UI) due to both taxation (8%) and NW labelling on HFSS food based on consumer response by QIMD (2026 – 2040)

**Figure 4.**
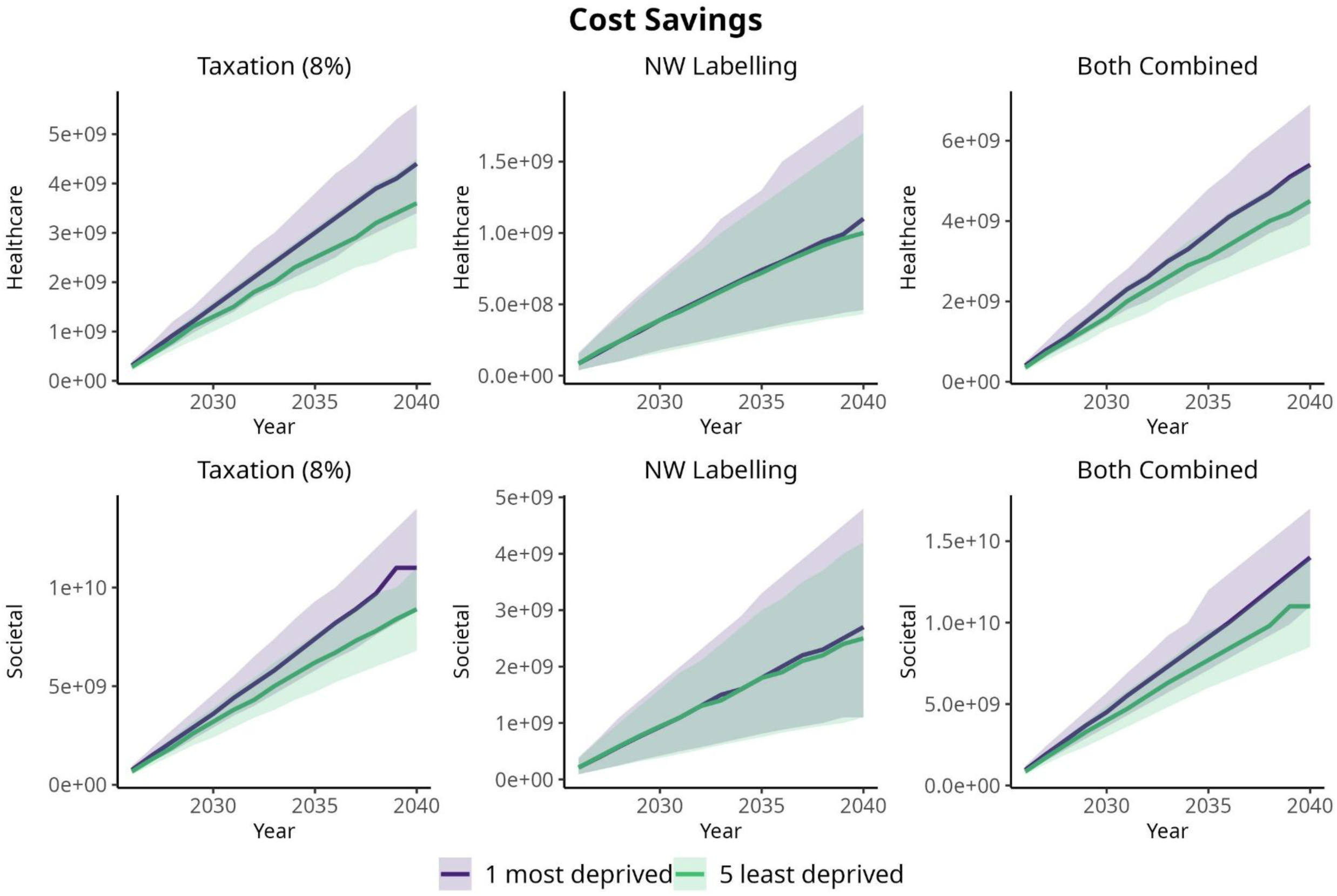
Cumulative cost savings (median; 95% UI) due to policy options based on consumer response by QIMD (2026 – 2040)

### Consumer response and reformulation

We assumed that product reformulation induced by the modelled policy would reduce the proportion of liable HFSS products subjected to either taxation or NW labelling. Findings from considering both consumer response and reformulation (Appendix Table 9) are similar to consumer response only (Table 2). Similar to the main analysis, more obesity, T2DM, and MLTC cases prevented, and QALYs gained were estimated for the most compared to least deprived areas (Appendix Figures 3, 4, and 5). We also projected greater cost savings in the most deprived groups (Appendix Figure 6). The absolute and relative equity slope indexes are similar to those reported in the main analysis (Appendix Table 10).

## Discussion

The present study offers the first modelling of the impacts of taxation, NW labelling, and both policies combined on HFSS products on a range of health outcomes, including CVDs, MLTCs, and related economic impact within the UK context. We estimated greater impacts would be achieved if taxation and NW labelling on HFSS products were implemented together. Under the assumption that the policies have the same effects across SES, both policies, separately and in combination, could reduce absolute and relative inequality as greater benefits were estimated in the most deprived areas.

Our findings are consistent with previous modelling studies on the impacts of taxation or NW labelling on unhealthy food. A modelling study from New Zealand supports our findings, demonstrating benefits from implementing Mexico’s 8% tax rate.^36^ The 8% tax on ‘junk food’ was projected to reduce DM incidence by 7%, CHD incidence by 3%, and the all-cause mortality rate by 1% over a 30-year simulation period, and to gain 561,000 health-adjusted life years over the population’s remaining lifetime. In England, a previous modelling study estimated a 2.64 percentage point reduction in obesity prevalence and 46 000 obesity-related deaths prevented if NW labelling were implemented on all liable packaged products over 20 years.^15^ Evidence from Mexico suggests that NW labelling could reduce 1.3 million obesity cases, or by 4.98 percentage points in its prevalence over five years.^37^

Our research highlights the benefits of implementing multiple measures, here taxation and NW labelling, on HFSS products to combat obesity in the UK. We also estimated equitable public health impacts from both policies, supported by a previous study on NW labelling in England.^15^ Taxation and NW labelling could be introduced as part of a broader set of policies targeting HFSS products, complementing existing measures (HFSS placement, advertising, and promotions), following Chile’s multi-policy strategy to address unhealthy foods, implemented since 2016.^38^ HFSS taxation and NW labelling could together reduce healthcare costs alone by £27 billion over 15 years (approximately £1.8 billion per year), equivalent to a 16% reduction in the current obesity-related NHS burden of £11.4 billion annually.^1^

The present study may be the first modelled evidence of food policy impact on multimorbidity or MLTCs. Given the growing challenge of MLTCs, with its prevalence predicted to increase by one-third from 2019 to 2049 in England,^39^ our findings also offer important insights for current prevention efforts. Our modelling with IMPACT_NCD_ was supported by high-quality data, including routine health records (e.g., CPRD Aurum, HES, ONS mortality records) calibrated with national estimates that increase confidence in the estimated policy impacts. We also conducted a range of sensitivity analyses that largely support the findings from the main analyses.

The present study has some limitations. We broadly classified foods into HFSS and non-HFSS based on the UK NPM. The assumed taxation effect on consumer response was derived from a consumer demand model specific to Scotland,^9^ which mainly defined HFSS based on discretionary foods. However, non-discretionary HFSS food groups also have own-price elasticities similar to discretionary ones (appendix p 6).^9^ Importantly, the assumed tax effect in the present study aligns with findings from a published meta-analysis^20^ and empirical data from post-taxation in Mexico.^10^ While a broad classification of HFSS may overlook cross-price elasticity and potential substitution effects, we also included the impact of HFSS food taxation on non-HFSS foods, such as a small decrease in fruit and vegetable intake in the main modelling scenario, as informed by the same demand model.^9^ We found similar impacts when reductions in fruits and vegetables were not accounted for (Appendix Table 5).

The tax effect on the intake of other non-taxed foods may be minimal, as empirical data from Mexico suggested no changes in sales of untaxed foods after the implementation of the 8% tax rate on unhealthy foods.^19^ Therefore, we assumed no compensation in the main analysis. However, we conducted a sensitivity analysis assuming minimal compensation of energy intake (11%) (Appendix Tables 11 and 12). This is based on evidence from a meta-analysis suggesting a non-significant energy intake compensation (11%) following reduced energy intake due to manipulations of food energy density.^40^ Future studies are now warranted to provide insights into the relevance of this assumption.

Assuming linearity in the policy effect, increasing the tax rate on HFSS products (e.g., doubling the tax rate to 16%) is likely to yield greater benefits, as demonstrated by a modelling study in Mexico.^22^ Substantially increasing the tax rate on all HFSS products may be important to avoid unhealthy food substitutions and reduce demand for these products.^41^ However, increased price of HFSS products may not only shift demand away from these items, but also affect the consumption of non-HFSS products, such as a potential further decrease in fruit and vegetable intake,^9^ which at present, has also been considered in our modelling. While changes in purchases of untaxed foods were not observed following the taxation in Mexico,^19^ the health benefits of increasing the HFSS tax rate may be partially offset by a decrease in the consumption of healthy products to some extent. Therefore, tax revenues could be used to subsidise fruits and vegetables, particularly for low SES groups who are likely to be disproportionately impacted by price increases.^41,42^

We used the effects of NW labelling from a meta-analysis by Song et al.^14^ for main (Table 2) and sensitivity analyses (Appendix Table 7). Positive findings from both strengthen the case for implementing NW labelling alongside taxation. However, large differences in the estimated impacts indicate the need for careful consideration when assuming policy effects. Although this is the most recent available evidence and was also used in a previous modelling study,^15^ it was primarily estimated from studies in controlled settings and outside the UK, and may not accurately reflect shopping behaviour in real-world environments. However, there is some evidence that warning labels on packaged products affect choices made by UK consumers.^43^ We assumed 25% of HFSS products without MLT labelling, based on the proportion of packaged products without MLT labelling,^15,25^ and did not consider possible different coverages by food groups due to limited data. The actual coverage of HFSS products without MLT labelling may be higher, as indicated by a previous study on fewer HFSS products with front-of-pack nutrient labelling (59%) compared to non-HFSS products (46%) sold within in-store restricted areas.^44^ Therefore, we may underestimate the policy impacts to some extent. Moreover, we did not consider energy compensation in response to NW labelling in the main analysis due to a lack of supporting direct evidence. Compensation would, however, be predicted to be minimal (11%) as studies suggest consumers are largely insensitive to changes in product energy density due to reductions in fat or sugar.^40^ A sensitivity analysis considering compensation (11%) is presented in Appendix Tables 11 and 12.

We assumed that household-level purchases were evenly distributed among individuals, and that potential differences in actual intake by sex and age are likely to be mitigated when comparing scenarios.^15^ Changes in purchases were assumed to reflect changes in intake without accounting for food waste.^15,22^ This is likely reasonable for non-out-of-home HFSS products, which are typically packaged and less perishable compared to fresh produce, and contribute to a small volume (16%, but high energy intake) of the total daily food weight (in grams).^6^ We assumed stable policy effects throughout the simulation period due to limited contrasting evidence. However, the assumed policy effect may change over time if individuals habituated to information, relevant policy campaigns, market shifts, policy amendments, and the introduction of new policy.^45^ Conversely, it is also plausible that policy effects could become somewhat larger over time (e.g., increased understanding and importance attached to warning labels after initial implementation). We used NDNS data from 2009 to 2019 to project trends in HFSS throughout the modelled horizon. This may not capture changes in dietary patterns resulting from the COVID-19 pandemic and recent economic downturns. We also simulated the policy effects in individuals aged 30-99 years, which did not account for impacts on children and younger adults. Nonetheless, this focus is relevant because middle-aged and older adults have a higher risk of developing the modelled health conditions.

We assumed similar effects of taxation and NW labelling across age, sex, and SES due to limited evidence. While this aligns with mixed findings or no evidence on differential effects reported by meta-analyses on sugary drink taxation^21^ and energy labelling^26^, low SES group may be more affected by taxation because they are generally more responsive to price increases.^41^ Compared to the main analysis, which assumed the same effect across income groups (Table 2), our sensitivity analysis on greater tax effects for lower income groups provided consistent and stronger evidence of its impact on reducing inequalities (Appendix Table 6).

We only estimated the policy effects on changes in energy and salt intake. We did not include pathways through reductions in sugar and total or saturated fat. Previous modelling studies assumed sugar and fat to operate through energy (e.g., 1 gram of sugar = 3.75 kcal),^4,46,47^ In addition, previous meta-analyses indicate less convincing evidence for the association between reduced dietary saturated or total fat and reduced CVD outcomes.^48–51^ A case-cohort study of nine European countries reported mixed findings on the link between dietary saturated fat and incident CHD, with the results varying by the food sources.^52^

As part of the sensitivity analyses, the potential reformulation driven by HFSS taxation was derived from a study on sugar and salt reduction targets with assumed coverage from other settings due to a lack of direct evidence.^28^ We estimated similar impacts when considering both consumer response and reformulation (vs. consumer response alone) because we assumed that reformulation decreased the liable products subjected to either taxation or NW labelling. Future studies are needed to test the relevance of this assumption. Moreover, sugar and salt reduction targets are currently existing UK policies, and these may lessen the reformulation effects assumed in the present study.

We did not consider some unintended consequences of HFSS taxation and NW labelling in our modelling. For example, evidence from an online survey in Hungary indicates adverse impacts of taxation on smaller producers, including financial loss and employment reduction.^41,53^ As the HFSS taxation and NW labelling would target key nutrients (e.g., sugar, salt), producers may respond by substituting them with artificial ingredients.^41^

The present study provides support for the UK Government to consider taxation and NW labelling as additional measures to address HFSS products and tackle obesity and diet-related diseases. The combined policies also potentially ease the current burden of obesity on the NHS,^1^ and both policies garner support from the public.^43,54^ A modest tax rate of 8% is particularly relevant for initial implementation in the UK alongside NW labelling, especially since other policies are already in place (e.g., HFSS placement, advertisement, and promotions). Considering the public response to its initial implementation, the tax rate could be gradually increased in line with existing policy practices for taxing ultra-processed food in Colombia^55^ and WHO’s recommendation of a 20% tax for SSBs.^56^ Importantly, implementation of taxation and NW labelling would benefit from public campaigns, as evidenced by the success of SDIL.^57^ This aligns with the literature on the importance of tax saliency (i.e., providing explicit information on pricing, tax implications) to enhance consumer awareness and taxation impacts in reducing sales of targeted products.^41,58^

## Conclusion

Implementing taxation and NW labelling on HFSS products could provide substantial and equitable public health benefits. These policies should be considered as additional measures to strengthen existing efforts to address obesity and diet-related diseases in the UK.

## Data Availability

The IMPACTNCD is an open-source modelling framework implemented in R, and the model and associated R scripts can be accessed at https://github.com/ChristK/IMPACTncd_Engl. For the present modelling study, additional R scripts are available at https://github.com/ediputra-ign/HFSS_taxation_and_NW_labelling. Detailed model description is available at https://www.health.org.uk/sites/default/files/2024-04/health_inequalities_in_2040_technical_appendix.pdf.

## Data sharing

The IMPACT_NCD_ is an open-source modelling framework implemented in R, and the model and associated R scripts can be accessed at https://github.com/ChristK/IMPACTncd_Engl. For the present modelling study, additional R scripts are available at https://github.com/ediputra-ign/HFSS_taxation_and_NW_labelling. Detailed model description is available at https://www.health.org.uk/sites/default/files/2024-04/health_inequalities_in_2040_technical_appendix.pdf.

## Acknowledgements

We thank Prof. Peter Scarborough, PhD, for providing early feedback on the modelling plans.

## Ethical approval

Ethical approval was not required for this study.

## Authors’ contributions

All authors (IGNEP, BC, ZC, RE, BA-C, ER, MO’F, and CK) contributed to the study design. CK developed and validated the IMPACT_NCD_ model. IGNEP, BC, CK accessed and verified the underlying data reported in this article. IGNEP conducted the analysis and wrote the original manuscript. All authors contributed to the data interpretation and revised the manuscript. All authors had final responsibility for the decision to submit for publication.

