## Supplementary appendix for "The estimated health and economic impacts of the introduction of taxation and warning labelling on foods high in fat, sugar or salt in England: A microsimulation study"

### Overview

We used the IMPACT$_{NCD}$, a validated dynamic, stochastic, discrete-time, open-cohort microsimulation model, to estimate the health and economic impacts of taxation and front-of-pack nutrient warning (NW) labelling on high in fat, sugar, or salt (HFSS) foods in England from 2026 to 2040. We compared the scenario of taxation with the counterfactual scenario of no taxation being implemented in England. For NW labelling (e.g., Appendix Figure 1A), we compared the scenario with voluntary implementation of multiple traffic light (MTL) labelling (e.g., Appendix Figure 1B) as the counterfactual scenario. In the final scenario, we estimated the impacts of both policies combined, assuming they are independent. We estimated the health impacts of the policies on cases prevented or postponed of obesity, cardiovascular diseases (CVDs) (a sum of coronary heart disease (CHD), stroke, atrial fibrillation, heart failure), type 2 diabetes mellitus (T2DM), multimorbidity or multiple long-term conditions (MLTCs), and all-cause mortality. Based on the perspective of economic evaluation, we estimated cost savings from healthcare (medical) and societal (a sum of medical, social care, informal care, and productivity) perspectives. We also calculate net monetary benefit, benefit-cost ratio, and incremental cost-effectiveness ratio to inform the cost-effectiveness of modelled policy scenarios.

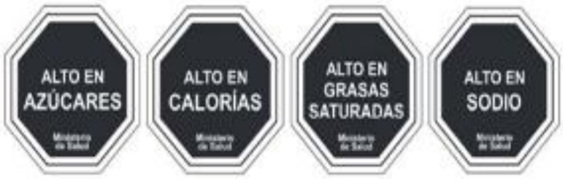

Notes: From left to right: High in Sugar, High in Calories, High in Saturated Fats and High in Sodium. At the bottom of each label it states Ministry of Health.

A

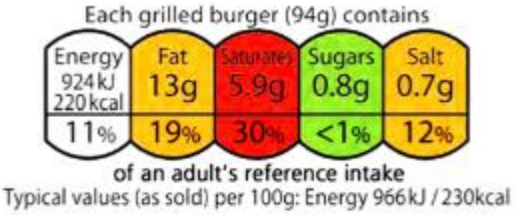

B

**Appendix Figure 1.** Nutrient warning (A) and multiple traffic light (B) labelling
Source: Department of Health and Social Care [1]

### Modelling approach and data sources

The IMPACT$_{NCD}$ was developed to understand the impacts of public health policies on non-communicable disease (NCD) prevention and has been used and validated in several countries, including England, Germany, the US, and Brazil.[2-5] For the present study, we used the IMPACT$_{NCD}$ developed for the English population. The IMPACT$_{NCD}$ is an open-source modelling framework implemented in R v4.5.1, and the main source code can be accessed at https://github.com/ChristK/IMPACTncd_Engl. For the present modelling study, additional codes or R scripts are available at https://github.com/ediputra-ign/HFSS_taxation_and_NW_labelling. The brief description of the IMPACT$_{NCD}$ in the present study follows the detailed model description available elsewhere.[6]

The close-to-reality synthetic population of England was created using data from different sources. The Office for National Statistics (ONS) population estimates and projections informed the population sizes over the simulation period. The Health Survey for England (HSE) informed the individual traits (sex, age, index of multiple deprivation (IMD), etc.) and trends in disease risk factors. These individual data linked to health care records (Clinical Practice Research Datalink (CPRD) Aurum linked to Hospital Episode Statistics (HES) and ONS mortality records) were used to inform trends in disease incidence, prevalence, and mortality. The epidemiological engine of IMPACT$_{NCD}$ consists of three modules: 1) sociodemographic (creating synthetic individuals from the joint age-sex distribution and assigning each individual to an IMD score and other relevant characteristics), 2) exposure (simulating life course exposure for each synthetic individual based on HSE data 2003 to 2014), and 3) disease modules (translating exposures into disease incidence using a population-attributable risk fraction (PARF) approach, taking into account for recovery and recurrence for some health conditions). Key assumptions for each module are available in the model description.[6] A flexible modelling approach, generalised additive models for location, shape, and scale (GAMLSS)[7,8] were used to predict trends in exposures, observed disease incidence and prevalence probability, and disease duration, based on sets of predictors (e.g., year, age, sex, ethnicity, IMD). GAMLSS can predict an assumed distribution of a dependent variable based on some function of independent variables. Compared to a linear regression that only models the mean value, GAMLSS accounts for both the mean and the standard deviation of a normally distributed dependent variable conditional on the independent variables.

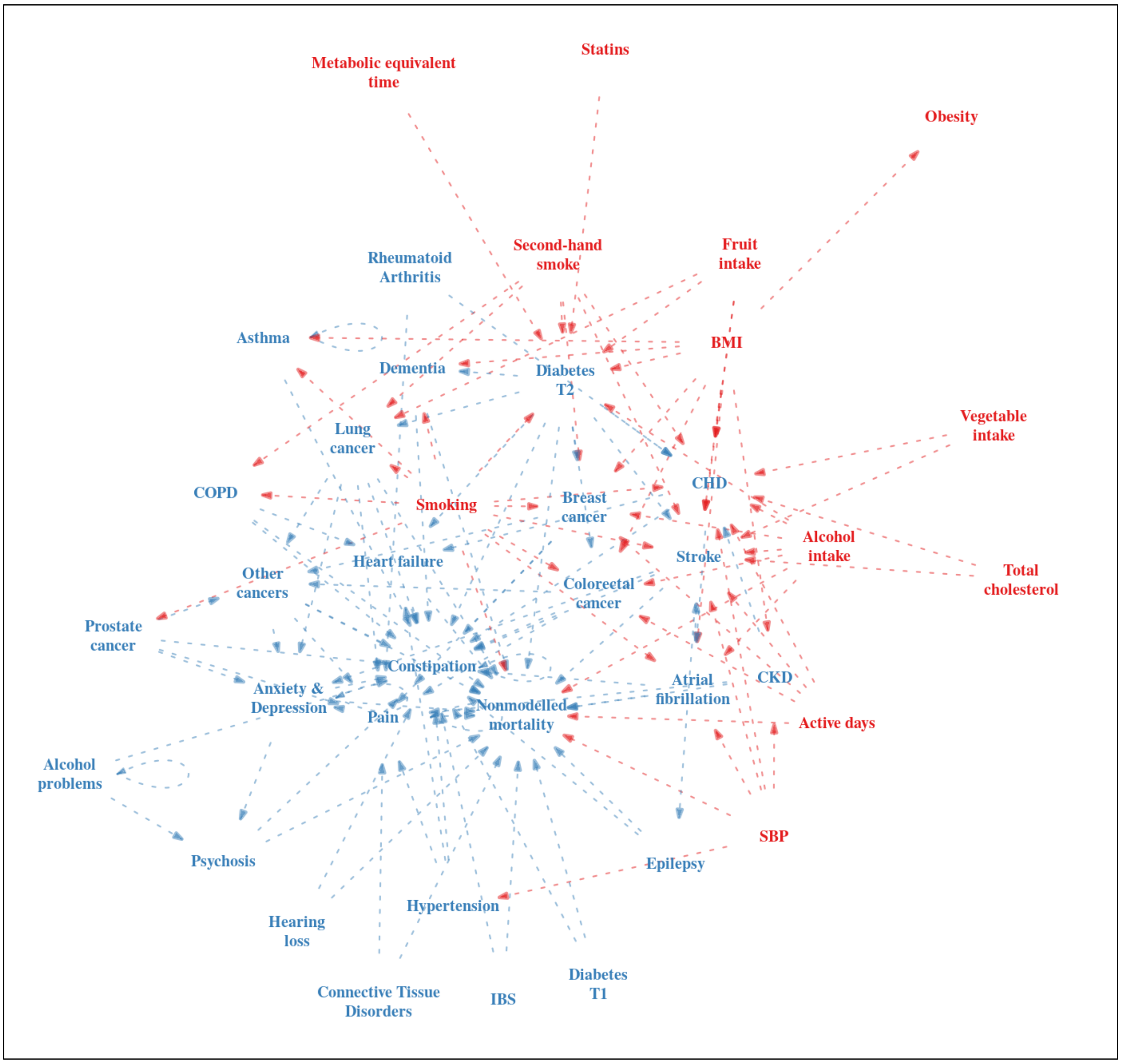

**Appendix Figure 2.** The causal diagrams of exposures and health outcomes in the IMPACT$_{NCD}$ model
Source: The IMPACT$_{NCD}$ model description[6]

The current updated IMPACT$_{NCD}$ includes eight main risk factors (smoking, body mass index (BMI), alcohol consumption, systolic blood pressure (SBP), total cholesterol, physical activity, fruit and vegetable consumption) modelled using HSE data and 20 health conditions (dementia, cancer, chronic obstructive pulmonary disease (COPD), atrial fibrillation, heart failure, constipation, chronic pain, epilepsy, stroke/transient ischaemic attack (TIA), type 1 or 2 DM, alcohol problems, psychosis/bipolar disorder, chronic kidney disease (CKD), anxiety/depression, CHD, connective tissue disorders, irritable bowel syndrome, asthma, hearing loss, and hypertension) modelled using CPRD Aurum (see the model description [6] on how each condition was defined). Based on these 20 health conditions, the Cambridge Multimorbidity Score (CMS) was calculated to assess the level of multimorbidity or MLTCs by assigning a weight or score based on the impacts of illness on the use of primary care, emergency health services, and risk of death.[6,9] We defined MLTCs as a CMS score > 1.5 (e.g., as in [10]).

The IMPACT$_{NCD}$ offers advanced microsimulation modelling and more complex and realistic modelling by incorporating competing causes of illness and death. For example, the model allows individuals to die from other modelled diseases when a particular policy may expand their lifespan due to reduced risk of death from a disease of interest. In addition, the $IMPACT_{NCD}$ models the policy impact on disease outcomes by considering concurrent trends in multiple risk factors, even when the policy primarily targets only one of them. Furthermore, the $IMPACT_{NCD}$ model structure was developed based on well-established causal pathways. The relationships between exposures (in red) and diseases or health conditions (in blue) are depicted in Appendix Figure 2. For better interpretation, the $IMPACT_{NCD}$ model description[6] provides detailed information and the structural diagrams for exposures affecting each modelled health condition.

### HFSS products: estimating energy and salt intake

We used the National Diet and Nutrition Survey (NDNS)[11] years 1 (2009) to 11 (2019) to estimate energy (kcal) and salt intake (grams) from HFSS products. Following a previous study on evaluating the contribution of HFSS products to the national diet,[12] we calculated per capita energy (kcal) and salt intake (grams) from HFSS based on the updated version of the UK nutrient profiling model (NPM) developed in 2018, which is likely to be implemented but has not yet been (originally developed in 2004/2005).[13] In brief, the UK NPM classified whether food falls into the HFSS category or not based on scoring the nutrient components. First, "A" points were calculated according to the amount of energy, free sugar, salt, and saturated fat. Second, the content of protein, fruit, fibre, vegetables and nuts contributed to "C" points. The final score was calculated by subtracting the "C" points from the "A" points. HFSS food with a score of ≥ 4 and beverages with a score of ≥ 1 were assigned to the HFSS category. We followed the codes provided by a recently published study in defining HFSS food or products.[12] This study also reported that the HFSS classification based on the UK NPM 2004/2005 and 2018 resulted in broadly similar results.[12] Based on the UK NPM 2018, HFSS products contribute to 41% and 42% of the per capita daily intake of energy and salt intake, respectively. We excluded the contribution of soft drinks as this type of beverage has been subject to another taxation in the UK (soft drink industry levy (SDIL)).[14]

Following the current implemented taxation on unhealthy food in supermarkets and retailers in Hungary[15] and Mexico,[16] we estimated the impact if the policy is implemented for all HFSS products from supermarkets and retailers (non-out-of-home (OOH) consumption; i.e., excluding food that is purchased and consumed from OOH food businesses, such as cafes, restaurants, etc.). First, we computed the percentage of OOH intake using NDNS data guided by a previous simulation modelling study estimating the impact of menu calorie labelling policy targeting OOH food businesses.[17] Two questions, "On average, how often do you/does child eat meals out in a restaurant or cafe?" and "On average, how often do you/does child eat take-away meals at home?" were used to calculate the proportion of intake from OOH. We assumed three meals a day, and the responses to the questions above were converted into the proportion as follows: "5 or more times per week" = 6/(7×3), "3-4 times per week" = 3.5/(7×3), "1-2 times per week" = 1.5/(7×3), "1-2 times per month" = 1.5/(30×3), "Rarely or never" = 0,[17] and then averaged to define the intake from OOH. We calculated the contribution of non-OOH HFSS products by multiplying energy and salt intake from HFSS food with the proportion of intake excluding out-of-home (OOH) (i.e., 1 – % of intake from OOH food businesses).

We used GAMLSS to model the trends in energy and salt intake from non-OOH HFSS food. GAMLSS created all the parameters of an assumed distribution of energy and salt intake and food weight, conditional on sets of predictors. We modelled trends in energy intake conditional on functions of year, sex, age, ethnicity, BMI, and IMD, and trends in salt intake conditional on functions of year, sex, age, ethnicity, and IMD. We also modelled the food weight (in grams) from HFSS products, which contribute to 16% of per capita food weight, using GAMLSS, conditional on year, sex, age, ethnicity, BMI, and IMD. All the parameters from the assumed distribution of the dependent variables, conditional on functions of the predictors, were applied to the population projections throughout the simulation period to predict the baseline distribution of energy and salt intake and food weight.

### Taxation on HFSS products

#### *Scenarios and coverage*

Our scenario for modelling taxation on HFSS products followed a real-world, currently implemented taxation rate of 8% on unhealthy food groups in Mexico.[18] This tax rate aligns with a relative tax rate ranging from 5% to 10% for most unhealthy products in Hungary.[19] This modelling scenario was compared to a counterfactual scenario of "no taxation" implemented on HFSS products in England.

We applied taxation to all HFSS products that account for approximately 41% and 42% per capita energy intake and salt intake, respectively (as the policy coverage). We excluded soft drinks and a corresponding proportion of HFSS products consumed from OOH food businesses (see "HFSS products: estimating energy and salt intake" above).

#### *Effect of taxation on HFSS products*

We estimated the effect of taxation mainly through consumer response (i.e., individuals reduce their energy and salt intake from HFSS products). We conducted a sensitivity analysis for the policy effect on reformulation (i.e., manufacturers reduce the content of nutrients of concern) due to limited evidence on this. We assumed that the effects remained consistent throughout the simulation period (2026-2040).

Consumer response to HFSS taxation

There are limited studies on a consumer-demand model assessing the price elasticity of HFSS products within the UK context. We used the best available evidence from Scotland that examined own- and cross-price elasticities of HFSS food groups (largely defined as discretionary food; classification was not based on the UK NPM).[20] The study found that a 10% tax (an increase in price of) on HFSS food groups may result in a 6-10% reduction in purchased HFSS products. Although a recent study examined price elasticities in the Scottish context, it does not provide a direct interpretation of the decrease in sales of overall HFSS products resulting from taxation (i.e., price increases).[21] Nevertheless, the recent study[21] reported price elasticities that were consistent with those used to inform the taxation effect in our modelling.[20] Importantly, the final assumed effect for an 8% tax rate is supported by previous studies (see below).

A recently published study by Viktorija, Yanaina [12] classifying HFSS products using the UK NPM suggested that "buns, cakes, pastries" (2.2%) (classified as HFSS in the Scottish study[20]) were among the top ten HFSS food groups that made the greatest contribution to the energy intake. Cheese (22.2%), butter (14.6%), sugars, preserves and sweet spreads (12.9%), and whole milk (8.6%) were among the other top ten HFSS products contributing to energy intake.[12] Even though these other categories were not mainly classified into HFSS products in a Scottish study,[20] their own-price elasticities were within a range of HFSS own-price elasticities (a 6-10% reduction in purchases due to a 10% price increase). For example, the own-price elasticity of dairy products, including cheese, butter, and whole milk, was −0.963, and the own-price elasticity of sugar and preserves was −0.872.[20] Therefore, we assumed that a 6-10% reduction in purchases due to a 10% tax rate (equivalent to 4.8 – 8% for an 8% tax rate) can be applied to a broader range of HFSS products based on the UK NPM definition. In addition, this assumed effect is similar to findings from a meta-analysis by Afshin, Peñalvo [22] suggesting that a 10% price increase reduced the consumption of unhealthy food (excluding SSB and fast food) by 9% (95% CI: [6, 12]). Similarly, empirical data from Mexico demonstrated that an 8% tax rate led to a reduction in sales of taxed food from 4.8% in the first year to 7.4% in the second year after the implementation.[23] Comparing the UK and Mexico, the nutritional quality of packaged products in the UK is slightly better than in Mexico (2.83 vs. 2.59; scores range from 0.5 to 5, with a higher score representing better quality).[24] Because food is classified differently in published studies, own-price elasticities are difficult to compare across all food groups between the two countries. However, the own-price elasticity for SSBs is similar between the UK (−0.98 to −1.17)[20,21] and Mexico (−1.16).[25] Similar own-price elasticities were also reported for fruits (−0.71 in the UK, −0.74 in Mexico) and meat (−0.75 in the UK, −0.72 to −0.87 in Mexico).[20,26] However, own-price elasticities for dairy products, vegetables, and prepared ready-to-eat foods were greater in Mexico than in the UK.[20,26] Although price elasticities differ between the UK and Mexico, the assumed taxation effect was informed by price elasticities derived from a consumer demand model within the UK context.

We applied the policy effect above (4.8 – 8% for an 8% tax rate) to baseline energy and salt intake from all HFSS products (excluding soft drinks and intake from OOH food businesses, see above). We used a uniform distribution to allow the taxation effect to range from 4.8 to 8%. We assumed that the quantity of products purchased is equal to their consumption. We also assumed the policy effect is similar across sociodemographic groups due to limited direct evidence in the UK, supported by high-quality evidence from sugary drink taxation.[27] Since we applied a proportional change to the baseline intake from HFSS products, which varied by sociodemographic characteristics, the actual change in intake would differ accordingly. We assumed a complete pass-through rate following a previous modelling study[28] and evidence of average pass-through rates of 105-108% for SDIL.[29]

Findings from the Scottish study also suggested that a 10% tax rate on HFSS products may lead to reductions in consumption of non-HFSS products, such as fruits and vegetables, by 2% and 5%, respectively.[20] Therefore, we included this in our modelling studies (equivalent reductions for an 8% tax rate are 1.6% and 4% for fruits and vegetables, respectively; 15% of uncertainties were added for the effects on fruits and vegetables).

Even though our assumption of no differential taxation effects by socioeconomic status (SES) is supported by a previous meta-analysis on sugary drink taxation,[27] the lower SES group may be more responsive towards price increases.[30] This is particularly evident in Mexico, where low-SES households exhibit a greater decrease in taxed food purchases.[18] Therefore, we conducted a sensitivity analysis using evidence from Mexico indicating a gradient in tax effects on reducing taxed food purchases by income levels: low (10.2%), middle (5.8%), and low (0%).[18] As we have five income categories, the effect for the group between low and middle income was taken as the average of those two groups, and similarly for the group between middle and high income.

Reformulation due to HFSS taxation (sensitivity analysis)

There is limited evidence on potential reformulation due to HFSS taxation. We used the best available evidence to make assumptions about reformulation. We drew evidence from the SDIL, which demonstrated an 11% reduction in sugar content per 100ml and a **6% reduction in calorie content** per serving for SDIL-targeted drinks during its announcement period (2015–2017).[31] In addition, the Institute for Fiscal Studies (IFS) calculated the likely reformulation due to potential taxation on added sugar and salt implemented on purchased food at and out-of-home, using Kantar data.[32] The reformulation was determined based on the maximum level of reformulation for each food category, assuming that manufacturers fully reformulated their products to meet the sugar and salt reduction targets. Based on this assumption, the potential reduction in sugar was estimated to range from 8.5 to 13 grams or 32 to 49 kcal (1 gram of sugar = 3.75 kcal).[32] Based on the mean daily energy intake for UK adults of 1700 to 1800 kcal[33] , with around 41%[12] (700 to 740 kcal) attributed to HFSS products (assuming this proportion is the same as the proportion of food subjected to sugar reduction targets calculated by the IFS), 32 to 49 kcal is equal to a 5% to 7% reduction. Based on all the evidence above, we assumed an average of **6% reformulation of the HFSS energy content**.

We also used evidence from the IFS for potential reformulation on salt, ranging from 0.6 to 0.7 grams.[32] Based on the average salt intake of 8.4 grams per day,[34] of which 54% (4.5 grams) was from food subjected to salt reduction targets,[35] 0.6 to 0.7 grams equate to a 13% to 16% reduction in salt intake. Therefore, we assumed **an average reformulation effect of 15% for salt in HFSS products**. The IFS calculated potential reformulation for energy and salt based on the sugar and salt reduction targets.[32] This is a limitation due to the different classifications between food targeted for sugar and salt taxation versus HFSS taxation.

*Coverage for reformulation.* There is limited evidence on the proportion of HFSS products that would be reformulated to avoid taxation. Evidence from Hungary indicates that 40% of the affected manufacturers modified their products to avoid taxation.[15] Therefore, assuming that these were equal to 40% of the HFSS products being reformulated, we applied the reformulation effect to 40% of the coverage above (e.g., reformulation coverage = 40% × non-OOH intake from HFSS).

*Adjustment for coverage due to consumer response and reformulation.* Reformulation may lead to a reduction of the proportion of taxed food as the manufacturers reduce the content of nutrients of concern. Therefore, we adjusted the coverage when estimating the combined effects of both consumer behaviour and reformulation (as part of the sensitivity analyses). Given that 40% of the HFSS products were assumed to be reformulated, the taxation would be applied to the remaining products (60%). For estimating the combined effects of consumer response and reformulation, the calculations are as follows.

Reduction in (energy/salt) intake = non-OOH (energy/salt) intake from HFSS × [**60% (without reformulation)** × (taxation effect) + **40% (reformulation)** × (reformulation effect)]

Post-intervention (energy/salt) intake = non-OOH (energy/salt) intake from HFSS × [1- (**60% (without reformulation)** × (taxation effect) + **40% (reformulation)** × (reformulation effect))]

### NW labelling on HFSS products

#### *Scenarios and coverage*

We implemented NW labelling on all non-OOH HFSS products (see "HFSS products: estimating energy and salt intake" above). This modelling scenario was compared to a counterfactual scenario of a currently implemented voluntary front-of-pack "MLT labelling" in England. A report suggests that 75% of packed products feature MTL labelling on their front of packs.[36] Therefore, we assumed that 75% of non-OOH HFSS products featuring MTL labelling in England do so. Even though 80% of all purchased products from supermarkets are packaged,[37] we assumed that almost all non-OOH HFSS products tend to be packaged because non-packaged fresh products (e.g., fruits, vegetables) are not HFSS and not subjected to NW labelling. In addition, NW labelling (e.g., "high in sugar") can also be added to the point-of-sale information or shelf-edge labels when HFSS products are not sold in packaging (e.g., fresh cake items in supermarkets). Therefore, the effect of NW labelling was implemented considering 75% of HFSS products featuring MTL labelling.

#### *Effect of NW labelling on HFSS products*

Similar to taxation, we also estimated the effect of NW labelling mainly through consumer response. We conducted a sensitivity analysis for the reformulation effect. We also assumed that the effects remained constant throughout the simulation period (2026-2040).

Consumer response to HFSS NW labelling

A network meta-analysis by Song et al.[38] suggests the effects of NW labelling (vs. control) in reducing total energy (kcal) purchased by 12.9% (95% CI: [8.0%; 17.9%]) and NW outperformed MTL labelling by 6.4% (95% CI: [0.4%; 12.5%]) in reducing total energy purchased. However, the same meta-analysis reported non-significant effects for i) NW labelling (vs. control) (7.8%; 95% CI: [-1.2%; 16.8%]) and ii) NW (vs. MTL labelling) (1.3%; 95% CI: [-7.7%; 10.3%]) in reducing total salt (grams). Song et al.[38] also reported significant effects of NW labelling on reducing energy purchased by 3.8% (95% CI: [0.9%, 6.8%]) per 100 grams of food. Still, they did not significantly differ when compared to MTL labelling in reducing total energy intake per 100 grams: 0.8% (95% CI: [-2.0%, 3.7%]). No significant effects were reported for NW labelling in reducing salt per 100 grams of food when compared to either control (2.1%; 95% CI: [-4.0%; 8.2%]) or MTL labelling (0.8%; 95% CI: [-4.9%; 6.5%]). Therefore, NW labelling may not outperform MTL labelling in reducing energy intake per 100-gram unit.

A meta-analysis by Croker et al.[39] reported an absolute reduction in energy and salt per 100 grams of food purchased. NW labelling (vs. control) reduced energy and sodium by 4.43 kcal (95% CI: [0.12, 8.74]) and 33.78 mg (95% CI: [8.16, 59.40]) per 100 grams of food or beverages purchased, respectively. Similar to the findings from Song et al.,[38] the effects of NW and MTL labelling on reducing energy and salt intake were similar in magnitude (further analysis was not carried out to examine differences), indicating that NW may outperform MTL labelling.

The two meta-analyses above consistently reported similar effects between NW and MTL labelling in reducing energy and salt intake per 100 grams of food purchased. Therefore, we used the effect sizes per 100 grams of food to inform our main modelling scenario. We decided to use the effect sizes reported as proportional changes (%) per 100 grams as reported by Song et al.[38] because these had been standardised across different baseline intakes from the included individual studies (as opposed to absolute changes from Croker et al.[39] that depend heavily on baseline values in the included studies). In terms of modelling, this would allow the absolute policy effects to change over time based on trends in baseline intake by sociodemographic characteristics. This is particularly important for modelling the policy effect on HFSS products that encompass a variety of food groups with differing energy contents and that are generally energy-dense (i.e., accounting for 41% of the per capita daily intake of energy (kcal), but only for 16% of the total food weight (grams)).[12] Furthermore, a meta-analysis by Song et al.[38] was published recently than Croker et al.[39] and includes a greater number of studies targeting multiple food types, which is often the case in the implementation of NW labelling targeting a wide range of products.

Following the effects from Song et al.,[38] we assumed that NW labelling would reduce energy by 3.8% (95% CI: [0.9%, 6.8%]) and salt by 2.1% (95% CI: [-4.0%, 8.2%]) per 100 grams of food purchased (assuming the quantity of food purchased equates to its consumption). As the effect sizes are per 100-gram unit, we calculated equivalent effects in reducing the total energy and salt intake by accounting for the total food weight (grams) consumed from HFSS products (see "HFSS products: estimating energy and salt intake"). As the effects of NW and MTL labelling were similar in reducing energy and salt intake, and 75% of packaged products feature MTL labelling,[36,40] we applied the policy effects to 25% of HFSS products not featuring MTL labelling. We did not include the reported estimates for NW vs. MTL labelling per 100 grams of food due to very small effects. We assumed the effects are similar across sociodemographic characteristics, following the current literature.[38-40] The actual coverage of HFSS products without MLT labelling may be higher than 25%, and we could be underestimating the policy impacts. A study reported that for all products sold within in-store restricted areas, front-of-pack nutrient labelling (FOPNL) was slightly higher in non-HFSS (59%) compared to HFSS products (46%).[41] Across products considered in-scope categories of placement restrictions (e.g., savoury snacks, breakfast cereals) sold within in-store restricted areas, a statistically higher proportion of non-HFSS products displayed FOPNL (59%) compared to those that were HFSS (18%).[41]

Following a previous simulation modelling study,[40] we conducted a sensitivity analysis using estimates from Song et al.[38] for reductions in total energy intake (as opposed to a reduction per 100 grams of food). We implemented a 12.9% (95% CI: [8.0%; 17.9%]) reduction in total energy to 25% of HFSS products without MTL labelling and 6.4% (95% CI: [0.4%; 12.5%]) to 75% of HFSS products with MTL labelling. Similarly, there were reductions in total salt intake of 7.8% (95% CI: [-1.2%; 16.8%]) for 75% of HFSS products and 1.3% (95% CI: [-7.7%; 10.3%]) for the remaining 25%. The effects of NW labelling on salt, as reported by Song et al.[38] (per 100 grams of food or total salt), are not statistically significant, suggesting that NW labelling may have no effect on salt intake. However, this is contradicted by a meta-analysis by Croker et al.[39] reporting a statistically significant impact of NW labelling on salt reduction (see above). In our simulation, whenever an opposite effect (i.e., an increase in salt intake due to NW labelling) was drawn from the random distribution, it was recoded as zero, indicating no effect.

<u>Reformulation due to HFSS NW labelling (sensitivity analysis)</u>
We conducted a sensitivity analysis for the reformulation effect. There is limited evidence from a meta-analysis for the reformulation effect specific to NW labelling. Findings from a meta-analysis on food labelling (including different types of labelling) reported a non-statistically significant effect of 0.9% (95% CI: [–3.1, 4.9]) for energy content reduction.[42] The same meta-analysis also reported that food labelling decreased sodium content by 8.9% (95% CI: [–17.3%, –0.6%]).[42] We used the reformulation effect above for NW labelling.

*Coverage for reformulation.* There is limited evidence on the proportion of HFSS products that would be formulated due to NW labelling. We used evidence from Chile suggesting that the proportion of products with NW labelling decreased by 20% due to potential reformulation.[43] We applied the reformulation effect to 20% of HFSS products.

*Adjustment for coverage due to consumer response and reformulation.* We assumed that reformulation also decreases the proportion of liable HFSS products subjected to NW labelling. We adjusted the coverage to model the combined effects of both consumer and reformulation effects (as part of the sensitivity analyses). Because 20% of the HFSS products were assumed to be reformulated due to NW labelling, we implemented the policy effect on changing consumer behaviour to the remaining products (80%). We did not limit the reformulation effect to only HFSS products not featuring MTL (25%) because evidence indicates that mandatory policies of front-of-package labelling are likely to have effects on product reformulation compared to voluntary approaches.[44] For estimating the combined effects of consumer response and reformulation, the calculations are as follows.

Reduction in (energy/salt) intake = non-OOH (energy/salt) intake from HFSS × [**80% (without reformulation)** × (25% for HFSS not featuring MTL labelling) × NW labelling effect per 100 grams of food) + **20% (reformulation)** × (reformulation effect)]

Post-intervention (energy/salt) intake = non-OOH (energy/salt) intake from HFSS – reduction (as above)

*(For sensitivity analysis, the coverage 75% for HFSS featuring MTL labelling was included)*

### The combined effects of taxation and NW labelling on HFSS products

Findings from four experimental studies (laboratory,[45] marketplace,[46,47] virtual grocery store settings[48]) indicate consistent evidence of no interactions between taxation and nutrition labelling in influencing diet outcomes (e.g., energy intake) in adults. Therefore, we assumed that both policies are independent, and we modelled the simultaneous effects of the policies using a multiplicative approach. Following the SimSmoke tobacco control policy simulation model approach used to estimate the impact of different policies combined (e.g., taxation, health warning), "the effect of a second policy is reduced by (1-the effect of the first policy) if another policy is simultaneously implemented".[49] The effect of combined policies i and j ("the relative effect of a policy is independent of other policies in effect" and "the effects are assumed to be multiplicative through their respective percentage reductions") is "(1+PRi) × (1+PRj)" with PR as a "per cent reduction" <0.[50,51] For example, the 8% tax rate with an assumed effect of -5% in reducing energy intake would result in post-intervention energy intake = baseline intake × 95%. Similarly, the effect of NW labelling from Song et al[38] (-4%) would result in post-intervention energy intake = baseline intake × 96%. For both policies combined, post-intervention energy intake = baseline intake × 95% × 96% = baseline intake × 91.2%. The overall effect of the policies combined in reducing energy intake is 8.8%, and the calculation can be simplified as 1 – [(1+PRi) × (1+PRj)].

Due to differences in coverage for taxation and NW labelling in reducing energy and salt intake, the post-intervention energy intake for consumer response is as follows. Due to larger coverage for HFSS taxation, we applied the effect of NW labelling to the post-intervention intake after taxation.

Post-intervention intake after taxation only (**A**) = non-OOH HFSS intake × (1-taxation effect)

Post-intervention intake after taxation and NW labelling (**B**) = **A** × 0.75 + **A** × (25% for HFSS not featuring MTL labelling) × (1-NW labelling effect per 100 grams of food)

The post-intervention intake accounting for both consumer response and reformulation was calculated as follows. We included reformulation for taxation only (40%) due to a larger coverage than the reformulation of NW labelling. The coverage for taxation-driven reformulation (40%) would account for potential reformulation due to NW labelling (20%). We also assumed that reformulation due to taxation may occur more quickly than NW labelling.

Post-intervention intake after taxation and NW labelling (**C**) = **B** × 60% + non-OOH HFSS intake × 40% × (1-reformulation effect due to taxation)

### Calorie and salt compensation and substitutions

For the main analysis, we did not account for calorie or salt compensation (i.e., reduced intake being partially offset by more intake later in the day) and substitutions for NW labelling due to limited evidence. For HFSS taxation, we included the potential impact of the policy on non-HFSS products, such as fruits and vegetables (see "Consumer response to HFSS taxation"). Evidence from Mexico suggests that while the sales of taxed food decreased, no changes in purchases of untaxed food were reported after implementing an 8% tax rate on unhealthy food.[18,28] Nevertheless, a previous meta-analysis study suggested minimal energy intake compensation due to food energy density manipulations (11%).[52] Therefore, we conducted an additional sensitivity analysis assuming energy intake compensation of 11% for both taxation and NW labelling. However, we did not consider salt compensation as supported by previous studies. Bolhuis et al.[53] reported that salt reduction in bread (with and without flavour compensation, e.g., potassium chloride and yeast extract) was not associated with sodium intake compensation. A randomised controlled trial in an experimental real-life canteen setting found that consumption of reduced-sodium foods did not lead to compensation behaviour.[54]

### Estimating the effect of changes in energy and salt intake

We assumed that a change in energy and salt intake would immediately impact BMI and SBP.

#### *Estimating the effect of a change in energy intake on BMI*

We used the "bw" package[55] that implements dynamic weight change mathematical models from Hall, Butte [56] and Chow and Hall [57] to estimate the impact of a reduction in energy intake on the change in BMI. These models estimate how body weight changes over time based on the principle of energy balance, which is determined by the difference between energy intake and expenditure.[46] Weight change-related physiological processes were modelled, accounting for each of the biological mechanisms involved. To calculate the change in BMI in adults, information on age, sex, weight (kg), height (m), and energy reduction per day from baseline energy intake was used as the model input.[55]

We also added the subsequent effect of a change in BMI on SBP and total cholesterol. We used evidence from a meta-analysis reporting SBP reduction by 5.79 mmHg (95% CI: [3.54, 8.05]) after a mean BMI reduction of 2.27 kg/m$^2$ in individuals with baseline BMI ≥ 25 kg/m$^2$.[58] Following pharmacologic interventions, per 1 kg/m$^2$ reduction in mean BMI, total cholesterol decreased by 8.87 (95% CI: [5.94, 11.79]) mg/dL or 0.229 mmol/L (95% CI: [0.154, 0.305]) in individuals with baseline BMI ≥ 25 kg/m$^2$.[59]

#### *Estimating the effect of a change in salt intake on SBP*

We used a meta-regression equation reported in a meta-analysis of 103 trials by Mozaffarian, Fahimi [60] to estimate the change in SBP (mmHg) due to a reduction in salt intake (grams). The meta-regression equation was "Y = α + B1X1 + B2X2 + B3X3", where Y = change in SBP (mmHg) per a reduction in sodium by 100 mmol/day (2.3 grams/day sodium; 5.85 grams/day salt); α = -3.735 (0.730), a constant, representing the effect of a 100 mmol/d sodium reduction on SBP in non-Black individuals with age =50 and without hypertension, B1 = -0.105 (0.029), for age centred at 50; B2 = -1.874 (0.884) for hypertensive status; and B3 = -2.489 (1.188 ) for Black ethnicity. The meta-analysis also suggested that a mean sodium intake of 1500 mg/day (3.75 grams of salt) was associated with a better outcome (i.e., reduced SBP) in randomised controlled trials. Therefore, we assumed that there was no policy effect on SBP if individuals consumed ≤ 3.75 grams of salt at baseline.

### Estimating the effect of changes in BMI and SBP on the disease outcomes

Appendix Figure 1 and the IMPACT$_{NCD}$ model description[6] show diseases that would be affected by changes in BMI, SBP, and other risk factors. In brief, to estimate the change in a risk factor (BMI, SMP) on the change in risk of disease incidence, the IMPACT$_{NCD}$ used relative risk (RR) estimates from published high-quality meta-analyses and Global Burden of Diseases (GBD) meta-analyses. The IMPACT$_{NCD}$ assumed multiplicative risk effects that combined the impact of changes in multiple risk factors on the probability of developing a modelled disease. The disease module in the IMPACT$_{NCD}$ description[6] provides details on RRs used for each modelled association between an exposure and a disease or health condition (pages 37 to 137 in the model description[6]). The IMPACT$_{NCD}$ assumed different lag times between exposure and outcome (4-5 years for most pairs of exposures and outcomes, except a mean lag time of 9 years for cancers), as changes in exposures or risk factors would not immediately impact changes in the disease incidence risk.

The IMPACT$_{NCD}$ calculated the population-attributable risk fraction (PARF), representing the proportion of the disease attributable to a risk factor, which depends on the RR and the prevalence of the risk factor in the population. Assuming multiplicative risk factors and exposures to risk factors are known at the individual level, PARF in the context of microsimulation modelling can be computed as follows (see [6] for detailed information).

$$PARF = 1 - \frac{n}{\sum_{i=1}^{n}(RR_{i1} * RR_{i2} * \dots * RR_{ik})} \quad [\mathbf{1}]$$

, with $n$ representing the total number of (synthetic) individuals and $RR_{i1...ik}$ is the unique individual relative risk of the disease associated with modelled risk factors. PARF was calculated stratified by some sociodemographic characteristics, including age, sex, ethnicity, and IMD. Depending on the disease, the $IMPACT_{NCD}$ assigned an RR of 1 if the level of risk factors does not increase the risk of the disease. For example, BMI < 22 kg/m$^2$, SBP < 115 mmHg, total cholesterol < 3.8 mmol/l, etc., were considered to have a RR of 1 for CHD (see [6] for detailed information on optimal levels for other risk factor-and-disease pairs).

The disease incidence not attributable to the risk factor was calculated using the following formula.

$$I_{Theoretical\ minimum} = I_{Observed} \text{ x } (1 - PARF) \quad [\mathbf{2}]$$

$I_{Observed}$ is the observed disease incidence, and $I_{Theoretical\ minimum}$ refers to the estimated disease incidence if the risk factor is optimal. $I_{Theoretical\ minimum}$ was calculated by age, sex, and IMD in the initial year of simulation and assumed to be stable throughout the simulation period.

The $I_{Theoretical\ minimum}$ is the annual baseline probability of individuals developing a disease specific to their sociodemographic characteristics due to non-modelled risk factors. The individualised annual probability of developing a disease due to their risk factors is as follows.

$$P(CVD|age, sex, ethncity, IMD, exposures) = I_{Theoretical\ minimum} \text{ x } (RR_{i1} * RR_{i2} * ... * RR_{ik}) \quad [\mathbf{3}]$$

The $IMPACT_{NCD}$ also accounts for conditions with recovery and recurrence, dependencies between conditions, and disease duration (see [6] for detailed information). Using data from CPRD linked to HES and ONS mortality records, case fatality rates were calculated. Mortality from some conditions (e.g., hearing loss, anxiety and depression, and hypertension) was assumed to be 0 due to very low case fatality rates. GAMLSS was used to calculate the case fatality rates for other conditions conditional on year, age, sex, and IMD. All-cause mortality rates in the CPRD-linked dataset were lower than those reported by the ONS. To address this, mortality projections over the simulation period were developed using ONS mortality rates by single year of age from 2001 to 2019. A calibration factor was then applied to adjust the CPRD-derived mortality rates to align with national estimates.[6]

### Quality-adjusted life years (QALYs)

We calculated QALYs by multiplying the predicted years of life lived by individualised disease-specific utility weights following previous studies.[14,61] First, age-specific utility values based on the EuroQol 5-Dimension scale (EQ-5D) for the English population were derived from [62]. Individualised disease-specific utility weights were then calculated by adjusting the utility values from Janssen and Szende [62] with disutility values (decrement) based on health conditions modelled in $IMPACT_{NCD}$ and relevant sociodemographic characteristics from Sullivan, Slejko [63]. We applied a discount rate of 3.5% per annum from 2026 to QALYs based on guidance from the UK Treasury,[64] following a previous study.[61]

### Cost savings

We calculated costs from healthcare (i.e., medical or healthcare sector) and societal (i.e., a sum of medical, social care, informal care, and productivity) perspectives. Cost savings were calculated by subtracting the total costs under the intervention or policy scenario from the total corresponding costs under the counterfactual scenario. We inflated the costs to represent 2026 values using the UK Treasury gross domestic product (GDP) deflators.[65] A discount rate of 3.5% per annum was applied to all costs from 2026.[61,66,67]

#### *Medical or healthcare costs*

Costs from the formal health (medical) sector were mostly extracted from published studies or reports within the UK context and expressed as cost per case per year as follows.

**Appendix Table 1.** Health care costs

| Diseases | Healthcare costs (2026 values) | Studies |
|---|---|---|
| Without modelled diseases | £1,254 | Office for Budget Responsibility [68]: We used healthcare spending costs at age 18, assuming the prevalence of modelled diseases is very low among this population (£1,093; costing year in 2023) |
| Obesity | £1,836 | Pearson-Stuttard, Holloway [69]: The weighted average cost of individuals with obesity, but without the presence of obesity-related complications (£1,415; costing year in 2019). |
| Hypertension | £124 | Sheppard, Fletcher [70]: The average cost of treatment delivered by different health professionals (£86; costing year in 2012). |
| Type 1 diabetes | £4,756 | Hex, MacDonald [71]: The weighted average cost excluding treatments for complications (£3,938; costing year in 2021) |
| Type 2 diabetes | £1,116 | Hex, MacDonald [71]: The weighted average cost excluding treatments for complications (£924; costing year in 2021) |
| Coronary heart disease | £5,444 | Landeiro, Harris [72]: The cost per case/patient (£4,116; costing year in 2018) |
| Atrial fibrillation | £1,680 | Burdett and Lip [73]: The cost per case/patient (£1,322; costing year in 2020) |
| Heart failure | £8,233 | Hollingworth, Biswas [74]: The annual cost after HF diagnosis (£5,756; costing year in 2013) |
| Stroke | £8,673 | Landeiro, Harris [72]: The cost per case/patient (£6,558; costing year in 2018) |
| Dementia | £2,985 | Landeiro, Harris [72]: The cost per case/patient (£2,257; costing year in 2018) |
| Lung cancer | £2,778 | Frontiers Economics [75]: The annual cost for lung cancer (£2,422; costing year in 2023) |
| Breast cancer | £2,956 | Frontiers Economics [75]: The annual cost for breast cancer (£2,577; costing year in 2023) |
| Colorectal cancer | £3,755 | Frontiers Economics [75]: The annual cost for colorectal cancer (£3,273; costing year in 2023) |
| Prostate cancer | £3,139 | Frontiers Economics [75]: The annual cost for other cancers (£2,737; costing year in 2023) |
| Other cancer | £3,139 | Frontiers Economics [75]: The annual cost for other cancers (£2,737; costing year in 2023) |
| Chronic kidney disease | £1,221 | Kerr, Bray [76]: The average annual cost per case/patient (£795; costing year in 2009) |
| Asthma | £282 | Mukherjee, Stoddart [77]: The cost per case/patient (£190; costing year in 2011) |
| Chronic obstructive pulmonary disease | £2,344 | McLean, Hoogendoorn [78]: The cost per case/patient (£1,579; costing year in 2011) |
| Epilepsy | £530 | Cockerell, Hart [79]: The cost per case/patient (£236; costing year in 1992) |
| Hearing loss | £953 | McDaid, Park [80]: We used evidence from the Netherlands ($1,115; assuming 1$ = £0.75, and adjusted for the difference in healthcare spending per capita between the UK vs. the Netherlands from Papanicolas, Mossialos [81]) (£620; costing year in 2009). The result is similar using an estimate for the US,[80] adjusted for the difference in healthcare spending per capita between the UK vs. the US. |
| Anxiety and depression | £1,348 | McCrone, Dhanasiri [82]: The weighted average cost per case/patient with anxiety or depression (£830; costing year in 2007) |
| Psychosis | £3,314 | Ride, Kasteridis [83]: The estimation of cost per case/patient without depression and comorbidity (£2,317; costing year in 2013) |

| | | |
|---|---|---|
| Alcohol problems | £1,074 | Institute of Alcohol Studies [84]: The total healthcare costs divided by the estimated number of people with high-risk drinking (£889; costing year in 2021) |
| Connective tissue disorders | £2,002 | Pinedo-Villanueva, Westbury [85]: The total healthcare cost in individuals with muscle weakness (£1,429; costing year in 2014) |
| Chronic pain | £795 | Oppong, Lewis [86]: The cost per case/patient (£612; costing year in 2019) |
| Rheumatoid arthritis | £1,192 | Ping-Hsuan, Claudia [87]: The cost per case/patient (£919; costing year in 2019) |
| Irritable bowel syndrome | £514 | Total healthcare cost per case/patient was summed from the primary (Akehurst, Brazier [88]) and secondary use (Soubieres, Wilson [89]), adjusted by the number of people using primary and secondary care from Wilson, Roberts [90] (£354; costing year in 2012) |
| Constipation | £479 | Dowden [91]: Using annual minimum cost for treatment per case/patient (£362; costing year in 2018) |

#### *Social care costs*

We followed a previous study in calculating social care costs.[92] First, average annual social care spending by single year of age was derived from the Office for Budget Responsibility (OBR),[68] and then multiplied by the 2021 single-age-specific population census data for England[93] to generate aggregate social care costs by age. Second, we aggregated total social care costs of four main diseases (cancer, CHD, dementia, and stroke) by age, from combining the annual social care costs of these diseases reported by Landeiro, Harris [72] and the number of potential individuals with corresponding diseases (the disease-specific prevalence from the GBD[94] multiplied by the 2021 census data). These four diseases were selected because of the availability of evidence on social care costs, and BMI (or obesity) is linked to these diseases (see Appendix Figure 1). We also found limited high-quality evidence for the social care costs of other modelled health outcomes. While the OBR estimated social care costs based on local-authority (public) spending only, the study by Landeiro, Harris [72] calculated all potential social care costs that may include both public- and private-funded social care. Therefore, we inflated the ORB's figures with the estimates from the National Audit Office,[95] which suggests that private social care spending is 1.2 times that of public spending. We calculated the average age-specific social care costs without the contribution of the four diseases by subtracting disease-specific social care costs from the total OBR's inflated social care costs, divided by the population from the 2021 census. The corresponding annual disease-specific social care costs were then added for individuals with any of these diseases throughout the simulation.

#### *Informal care costs*

Informal (unpaid) care costs were calculated, following previous studies.[92,96] First, we used a regression formula developed by Rowen, Dixon [97] to predict the number of days individuals need informal care based on the Health Outcomes Data Repository for people (n = 44,494 individuals) discharged from a hospital in Wales. The formula used the zero-inflated negative binomial with variable inflation to estimate the number of days of informal care out of 42 days using sociodemographic information (age, sex), comorbidity, primary diseases by ICD Chapter, and individualised disease-specific utility weights (non-discounted EQ-5D index score; see "Quality-adjusted life years (QALYs)" ) as model inputs. Using the formula, we calculated the number of days of informal care per person per year specific to the characteristic above. We assumed 21 hours per week, or 3 hours per day, of receiving adult informal care, informed by a previous study using HSE data.[98] The average cost of informal care per hour was valued at £7.85 (2018 costing year, equivalent to £10.4 in 2026) based on the average hourly rate of a home care assistant,[99] following a previous study.[72] The annual informal care costs were computed by combining information on i) the number of days of informal care per year, ii) the average number of hours of informal care received per day, and iii) the hourly cost of informal care.

#### *Productivity costs*

We defined productivity costs following a previous approach.[92] Productivity consisted of i) paid and ii) unpaid work or production. The paid production was calculated based on estimates from the Annual Survey of Hours and Earnings (ASHE) on average paid hours worked and hourly earnings (inflated to 2026 value) by region, age, and sex.[100] These were then adjusted by population estimates by region, age, and sex,[101] and the employment rates by age and sex,[102] to estimate the weighted average annual earnings in England. For the unpaid production, we applied the regression formula from Claxton et al.[103] to calculate annual hours of unpaid production given sex and age. We assumed that the unpaid production is constant from ages 70 to 100.[92] Unpaid work was valued at £10 per hour (2015 costing year, equivalent to £13.8 in 2026) based on data from the ONS, suggesting, on average, women and men do 26- and 16-hour unpaid work a week, and they would earn £259.63 and £166.63 more per week if their unpaid work were paid, respectively.[104] We combined the annual hours of unpaid production and its hourly rate (£13.8) to estimate the annual monetary value of the unpaid production. Both annual paid and unpaid production constituted total productivity.

Claxton et al.[103] developed a formula to calculate productivity rate based on age and non-discounted EQ-5D index score. The full health productivity rate with EQ-5D = 1 ranges from 0% to 76%. We assumed individuals in perfect health condition (EQ-5D = 1) would have a 100% productivity rate, and therefore, we inflated the productivity rate to 100% for individuals with EQ-5D = 1. This was done by estimating a relative productivity, dividing the productivity rate of any EQ-5D index score by the full health productivity rate (EQ-5D = 1), following a previous approach.[92] The final total productivity was defined by multiplying the total productivity by the relative productivity. It is important to note that the total hours of unpaid production produced using the formula from Claxton et al.[103] may include time spent on unpaid adult care, and therefore, there may be some double counting when looking at the results from the societal perspective. However, this tends to be minimal as individuals in the UK spend less than 4 minutes per day on average for unpaid adult care.[105]

### Policy costs

We primarily considered UK Government policy impact assessments in calculating possible policy costs of HFSS taxation and NW labelling. The costs were inflated to represent 2026 values using the GDP deflators.[65] We also applied a discount rate of 3.5% per annum from 2026 to all policy costs.[61,66,67]

#### *Government administration, familiarisation, and enforcement costs*

There is no direct estimation of the government administration cost for a new policy calculated in three different impact assessments that have been done in the UK: the Food Standards Agency's 2009 Food Labelling Impact Assessment,[106] Department of Health and Social Care's 2020 Mandatory Calorie Labelling Impact Assessment,[107] and the Department of Health and Social Care's 2020 Restricting Volume Promotions for HFSS Products Impact Assessment.[108] A study from the US assumed $9,436,507 (2017 costing year) as the administrative cost for implementing added sugar labelling.[109] Within the UK context, Nesta commissioned HealthLumen to estimate the first-year (one-off) government implementation and familiarisation costs of sugar and salt taxation, which were £18 million (2024 costing year, equivalent to £19,316,600 in 2026, rounded to the nearest hundred).[110] Therefore, we assumed similar one-off total government administrative and familiarisation costs of £19,316,600 for HFSS taxation and NW labelling.

We assumed an enforcement cost of £68,000 per year (2020 costing year, equivalent to £86,395 in 2026), informed by the Department of Health and Social Care's 2020 Restricting Volume Promotions for HFSS Products Impact Assessment.[108] This was based on the duration and number of visits to relevant retail stores in England, multiplied by the hourly rate of a staff member for enforcement.[108] We assumed a similar annual monitoring cost for taxation and NW labelling on HFSS. Based on an enforcement cost of £86,395 per year, we estimated the monitoring cost for 15 years (throughout the simulation period).

#### *Industry familiarisation, assessment, and compliance costs*

Based on the Department of Health and Social Care's 2020 Restricting Volume Promotions for HFSS Products Impact Assessment,[108] we assumed a similar familiarisation cost for HFSS taxation and NW labelling of £0.2 million (2020 costing year, equivalent to £254,100 in 2026, rounded to the nearest hundred). The familiarisation cost assumed that it would take one manager-level employee one hour to read and become familiar with the policy. The hourly salary was then multiplied by the number of businesses in England (n = 557).[108]

We calculated the HFSS assessment cost as the industries or manufacturers would need to calculate the NPM scores. Following the Department of Health and Social Care's 2020 Restricting Volume Promotions for HFSS

Products Impact Assessment,[108] a half hour would be needed to assess one product and record the outcome. Assuming the hourly staff cost of £27.30 (including a 30% uplift) (2019 costing year, equivalent to £35.4 in 2026),[108] a half-hour assessment would cost £17.7 in 2026. One of the big four supermarkets, such as Tesco, sells 25,000 food and beverage product lines.[108,111] We assumed 10 main grocery supermarkets contributed to the top market shares,[112] and therefore, 250,000 product lines would need to be assessed. We added 50,000 product lines, possibly contributed by other small stores, making a total of 300,000 product lines. The Department of Health and Social Care's 2020 Restricting Volume Promotions for HFSS Products Impact Assessment[108] suggests that other big supermarkets have fewer products (e.g., Asda, Morrisons), and stores that are not supermarkets stock 1,000 products. Therefore, the total number of products above tends to be an overestimate. We estimated the one-off assessment cost of £5,313,700 (rounded to the nearest hundred), assumed to be similar for HFFS taxation and NW warning.

For taxation, we assumed no main direct potential costs for industries other than HFSS assessment and familiarisation costs above, following cost estimations for sugar and salt taxation[110] and the extension of SDIL to flavoured milks.[111] For NW labelling, we assumed that the food manufacturing industries would bear the cost of redesigning and reprinting the label. We followed a previous study by Collins et al.[66] and the Food Standards Agency's 2009 Food Labelling Impact Assessment,[106] assuming an average labelling cost of £1,000 (2009 costing year, equivalent to £1,536 in 2026) per Stock Keeping Unit (SKU) ("a food product with its own unique bar code"). As there is no data on the number of product lines targeted for NW labelling, we followed an approach used by Collins et al.[66] on modelling the costs of implementing front-of-pack labelling to reduce salt intake. Collins et al.[66] implemented the labelling cost (£1,000) to 20,000 product lines (assumed a total labelling cost of £20,000,000), which was based on the Food Standards Agency's consultation with the UK retailers and other stakeholders on "a best estimate of the number of lines reformulated within the processed foods categories".[66] To avoid underestimation, we applied it to 100,000 product lines, five times the size of the 20,000 product lines assumed in Collin et al.,[66] as this study focused on salt rather than the broader category of HFSS.[66] A big four supermarket, such as Tesco, stocks 25,000 food and beverage product lines.[108,111] However, not all would be affected by the policy, as recent evidence indicates that only 30% of consumed food items are classified as HFSS[12], and 30% of supermarket products are HFSS.[108] Therefore, 7,500 (30% x 25,000) product lines would be affected by the policy. This aligns with the Department of Health and Social Care's 2020 Restricting Volume Promotions for HFSS Products Impact Assessment[108], which assumed 7,500 products would be subjected to HFSS regulation in Tesco. We also assumed 10 main grocery supermarkets contributed to the top market shares,[112] and therefore, 75,000 product lines would fall within the policy scope. We added 25,000 product lines, possibly contributed by other small stores, making a total of 100,000 product lines. Therefore, we estimated a total one-time labelling cost of £153,614,200 (rounded to the nearest hundred). This tends to be an overestimate as discounters/small stores may have fewer product lines.[108]

***Industry reformulation costs (if the manufacturers choose to reformulate their products, for a sensitivity analysis only)***

There is limited data on the reformulation cost of food products. In the US, the reformulation cost ranges from $5,000 to $4,000,000 per product. According to Collins et al.,[66] the potential cost for reformulation was £25,000 (2009 costing year, equivalent to £38,404 in 2026) per product line, supplied by the British Retail Consortium. Following the modelling scenarios with reformulation (sensitivity analyses), we assumed that reformulation would occur in 40% and 20% of the products for taxation and NW labelling, respectively (see "Effect of taxation on HFSS products" and see "Effect of NW labelling on HFSS products"). Given 100,000 product lines assumed to be targeted by the policy, the reformulation cost would be implemented for 40,000 and 20,000 product lines for taxation and NW labelling with one-off costs at £1,536,141,600 and £768,070,800 (rounded to the nearest hundred), respectively. It is important to note that many products can be reformulated within a natural product cycle, and therefore, the reformulation costs may be overestimated.

**Appendix Table 2.** Policy costs based on different scenarios

| Scenarios | Government costs | | Industry costs | | | | Total |
|---|---|---|---|---|---|---|---|
| | **Administrative and familiarisation (One-off)** | **Enforcement (Ongoing; 15 years)** | **Familiarisation (One-off)** | **Assessment (One-off)** | **Compliance for product labelling (One-off)** | **Reformulation (One-off)** | |
| **NON-DISCOUNTED COSTS** | | | | | | | |
| *Main analysis (consumer response)* | | | | | | | |
| Taxation | £19,316,600 | £1,295,900 | £254,100 | £5,313,700 | - | - | £26,180,300 |
| NW labelling | £19,316,600 | £1,295,900 | £254,100 | £5,313,700 | £153,614,200 | - | £179,794,500 |
| Taxation and NW labelling[a] | £38,633,200 | £2,591,800 | £508,200 | £5,313,700[b] | £153,614,200 | - | £200,661,100 |
| *Sensitivity analysis (consumer response and reformulation)* | | | | | | | |
| Taxation | £19,316,600 | £1,295,900 | £254,100 | £5,313,700 | - | £1,536,141,600 | £1,562,321,900 |
| NW labelling | £19,316,600 | £1,295,900 | £254,100 | £5,313,700 | £122,891,300[c] | £768,070,800 | £917,142,400 |
| Taxation and NW labelling[a] | £38,633,200 | £2,591,800 | £508,200 | £5,313,700[b] | £122,891,300[c] | £1,536,141,600[d] | £1,706,079,800 |
| **DISCOUNTED COSTS** | | | | | | | |
| *Main analysis (consumer response)* | | | | | | | |
| Taxation | £18,663,400 | £995,000 | £245,500 | £5,134,000 | - | - | £25,037,900 |
| NW labelling | £18,663,400 | £995,000 | £245,500 | £5,134,000 | £148,419,500 | - | £173,457,400 |
| Taxation and NW labelling[a] | £37,326,800 | £1,990,000 | £491,000 | £5,134,000[b] | £148,419,500 | - | £193,361,300 |
| *Sensitivity analysis (consumer response and reformulation)* | | | | | | | |
| Taxation | £18,663,400 | £995,000 | £245,500 | £5,134,000 | - | £1,484,194,800 | £1,509,232,700 |
| NW labelling | £18,663,400 | £995,000 | £245,500 | £5,134,000 | £118,735,600[c] | £742,097,400 | £885,870,900 |
| Taxation and NW labelling[a] | £37,326,800 | £1,990,000 | £491,000 | £5,134,000[b] | £118,735,600[c] | £1,484,194,800[d] | £1,647,872,200 |

NW = nutrient warning

[a]We assumed that the cost would double if both policies were implemented, unless otherwise specified.

[b]Both taxation and NW warning target the same HFSS products; therefore, the assessment would be one time only.

[c]We assumed the reformulation for NW labelling applied to 20% of the products, and this reduced the proportion of liable products for NW labelling by 20% (see "Effect of NW labelling on HFSS products").

[d]When both policies are implemented together, we only used the proportion of reformulation for taxation at 40% (see "The combined effects of taxation and NW labelling on HFSS products").

### Net monetary benefit

Net monetary benefits refer to "the value of an intervention in monetary terms" given a specified willingness-to-pay threshold for each unit of benefit.[113] We calculated the net monetary benefit of each of the modelled policy scenarios based on estimates of i) cumulative lifetime health (QALYs) and ii) cumulative net costs (differences between cost savings and policy costs) (e.g., as in [14]). We used different thresholds or values per QALY: i) National Institute for Health and Care Excellence (NICE)'s lower threshold of £20,000; ii) NICE's upper threshold of £30,000;[114] and iii) the UK Treasury Green Book recommendation of £70,000.[115]

### Benefit-cost ratio

The benefit-cost ratio (cost-benefit analysis) was calculated as cumulative benefits (cumulative sums of the lifetime health (QALYs) and cost savings) divided by cumulative policy costs (see above). We calculated the benefit-cost ratio using different values or thresholds per QALY (see "Net monetary benefit").

### Incremental cost-effectiveness ratio

Following previous simulation modelling studies,[3,61] we calculated the incremental cost-effectiveness ratio (cost-utility analysis), cost per QALY gained, as the ratio of cumulative net costs (difference between cost savings and policy costs) by cumulative QALYs gained.

### Estimating model uncertainty

The $IMPACT_{NCD}$ used the 2[nd]-order Monte Carlo approach[116,117] to estimate the uncertainty for each scenario.[6] A random sampling method was used in the Monte Carlo approach to generate random values for uncertain inputs (e.g., policy effects, incidence, prevalence, case fatality rates, RRs) according to their assumed probability distributions. Using a different set of randomised inputs, uncertainties in predictions were then quantified by repeatedly running the model (based on the number of iterations).[116,117] See the $IMPACT_{NCD}$ for detailed information on uncertainty.[6]

**Appendix Table 3.** Data sources used in the model

We summarised the information on data sources used in the $IMPACT_{NCD}$. Detailed information can be found in the $IMPACT_{NCD}$ model description[6]

| **Parameters** | **Outcome** | **Details/Sources** | **Differences by sociodemographic groups or predictors** | **Projection distribution of the mean** | **Uncertainty** |
|---|---|---|---|---|---|
| ***Population data*** | | | | | |
| Population size, composition, and projection | Population | Official National Statistics (ONS) population projections up to 2043 | Year, age, sex, ethnicity, deprivation, local authority, and strategic health authority (SHA) | - | - |
| Disease incidence, prevalence, and mortality | Trends in disease incidence, prevalence, and mortality | Clinical Practice Research Datalink (CPRD) Aurum linked to Hospital Episode Statistics (HES) and ONS mortality records) 2019 | Year, age, sex, and deprivation | GAMLSS | |
| ***Exposures and risk factors*** | | | | | |
| Body mass index (BMI) ($kg/m^2$) | BMI | Health Survey for England (HSE) 2003 to 2014 | Year, age, sex, ethnicity, deprivation, SHA, smoking status | GAMLSS | - |
| Systolic blood pressure (SBP) (mmHg) | SBP | HSE 2003 to 2014 | Year, age, sex, ethnicity, deprivation, SHA, smoking status | GAMLSS | - |
| Total serum cholesterol (mmol/L) | Total serum cholesterol | HSE 2003 to 2014 | Year, age, sex, ethnicity, deprivation, and SHA | GAMLSS | - |
| Alcohol intake (grams/day) | Alcohol intake | HSE 2003 to 2014 | Year, age, sex, ethnicity, deprivation, and SHA | GAMLSS | - |
| Smoking status (current smoker/ ex-smoker/ never-smoker) | Smoking status | HSE 2003 to 2014 | Year, age, sex, ethnicity, deprivation, and SHA | GAMLSS | - |
| Smoking duration (years of smoking) | Smoking duration | HSE 2003 to 2014 | Year, age, sex, ethnicity, deprivation, and SHA | GAMLSS | - |
| Smoking intensity (cigarettes per day) | Smoking intensity | HSE 2003 to 2014 | Year, age, sex, ethnicity, deprivation, and SHA | GAMLSS | - |
| Environmental tobacco exposure (yes, no) | Environmental tobacco exposure | HSE 2003 to 2014 | Year, age, sex, ethnicity, deprivation, and SHA | GAMLSS | - |
| Fruit consumption (portions/day) | Fruit consumption | HSE 2003 to 2014 | Year, age, sex, ethnicity, deprivation, and SHA | GAMLSS | - |

| | | | | | |
|---|---|---|---|---|---|
| Vegetable consumption (portions/day) | Vegetable consumption | HSE 2003 to 2014 | Year, age, sex, ethnicity, deprivation, and SHA | GAMLSS | - |
| Physical activity (active days per week) | Physical activity | HSE 2003 to 2014 | Year, age, sex, ethnicity, deprivation, and SHA | GAMLSS | - |
| Statin prescriptions (yes, no) | Statin prescriptions | HSE 2012 to 2014 | Year, age, sex, ethnicity, deprivation, SHA, cholesterol | GAMLSS | - |
| Energy intake | Energy intake | National Diet and Nutrition Survey (NDNS) 2009 to 2019 | Year, age, sex, ethnicity, deprivation, and BMI | GAMLSS | - |
| Salt intake | Salt intake | NDNS 2009 to 2019 | Year, age, sex, ethnicity, and deprivation | GAMLSS | - |
| Food weight (grams) | Food weight (grams) | NDNS 2009 to 2019 | Year, age, sex, ethnicity, deprivation, and BMI | GAMLSS | - |
| ***Policy effects*** | | | | | |
| Effect of taxation on consumer behaviour | Change in energy and salt intake, fruit, and vegetable intakes | A consumer demand model from a study within the Scottish context by Nneli et al.[20] | Energy and salt: 4.8% to 8% for an 8% tax rate (no differential effect)<br>Fruit: 1.6% for an 8% tax rate (no differential effect)<br>Vegetable: 4% for an 8% tax rate (no differential effect) | - | Energy and salt: 4.8% to 8% (uniform distribution)<br>Fruit and vegetable: Mean ± 15% uncertainty |
| Effect of nutrient warning labelling on consumer behaviour | Change in energy and salt intake | A meta-analysis by Song et al.[38] | Energy: 3.8% (95% CI: [0.9%, 6.8%]; based on two studies) per 100 grams of food (main analysis; no differential effect); 12.9% (95% CI: [8.0%; 17.9%]; three studies) in reducing total energy and outperforming multiple traffic light (MLT) labelling by 6.4% (95% CI: [0.4%; 12.5%]; one study) in reducing total energy (sensitivity analysis; no differential effect)<br>Salt: 2.1% (95% CI: [-4.0%; 8.2%]; two studies) per 100 grams of food (main analysis; no differential effect); 7.8% (95% CI: [-1.2%; 16.8%]; one study) in reducing total salt and outperforming MTL labelling by 1.3% (95% CI: [-7.7%; 10.3%]; one study) in reducing total salt (sensitivity analysis; no differential effect) | - | Mean ± standard deviation (SD) |
| Effect of taxation on reformulation | Change in energy and salt intake | Calculated based on findings from the Institute for Fiscal Studies (IFS)[32] | Energy: 6% (no differential effect)<br>Salt: 15% (no differential effect) | - | Mean ± 15% uncertainty |

| | | | | | |
|---|---|---|---|---|---|
| Effect of nutrient warning labelling on reformulation | Change in energy and salt intake | A meta-analysis by Shangguan et al.[42] | Energy: 0.9% (95% CI: [–3.1, 4.9]; four studies) (no differential effect)<br><br>Salt: by 8.9% (95% CI: [–17.3%, –0.6%]; four studies) (no differential effect) | - | Mean ± SD |
| Effect of change in energy intake on BMI | Change in BMI | Based on dynamic weight change mathematical models from Hall et al.[56,57] | Age, sex, weight (kg), height (m), and energy reduction per day from baseline energy intake were used as the model inputs (See "Estimating the effect of change in energy intake on BMI"). | - | - |
| Effect of change in salt intake on SBP | Change in SBP | A meta-regression equation reported in a meta-analysis by Mozaffarian et al.[60] | Age, sex, baseline hypertensive status, ethnicity, and reduction in salt per day from baseline salt intake were used as the model inputs (See "Estimating the effect of change in salt intake on SBP"). | - | Mean ± SD |
| Risk factors:<br><br>BMI, SBP, total cholesterol, alcohol, smoking, environmental tobacco exposure, fruit and vegetable consumption, statin prescriptions | Health conditions:<br><br>Coronary heart disease, stroke, breast cancer, colorectal cancer, lung cancer, prostate cancer, other cancer, chronic obstructive pulmonary disease, atrial fibrillation, heart failure, type 1, type 2 diabetes mellitus, chronic kidney disease, dementia, hypertension, asthma, alcohol problems, anxiety and depression, constipation, irritable bowel syndrome, chronic pain, psychosis, epilepsy, connective tissue disorders, hearing loss, non-modelled diseases | The relative risks between pairs of risk factors and health conditions and dependencies with other health conditions (see Appendix Figure 1) were extracted from high-quality meta-analyses, including Global Burden of Disease (GBD) meta-analyses.<br><br>See the $IMPACT_{NCD}$ model description **pages 37 to 137** for detailed information.[6] | The reported relative risks were adjusted for different sociodemographic characteristics and some risk factors, varied by meta-analyses.<br><br>See the $IMPACT_{NCD}$ model description **pages 37 to 137** for detailed information.[6] | - | Mean ± SD |

**Appendix Table 4.** Assumptions implemented in the model

We summarised the assumptions used in the present study. Detailed assumptions used in the $IMPACT_{NCD}$ can be found in the $IMPACT_{NCD}$ model description[6]

| Components | Assumptions |
|---|---|
| Population data | We do not consider social mobility. |
| | We do not consider migration explicitly. |
| Exposures | The surveys used to inform exposures are truly representative of the population. |
| | The quantity of purchases equates to its intake (consumption) without waste. |
| | Household-level purchases were evenly distributed among individuals from the same household. |
| | The proportion of packaged products without multiple traffic light (MLT) labelling corresponds to the proportion of non-out-of-home high in fat, sugar or salt (HFSS) products without MLT labelling. |
| | The reformulation reduces the proportion of HFSS products subjected to either taxation or nutrient warning (NW) labelling. |
| Effect estimates | Similar policy effects across sociodemographic characteristics (age, sex, deprivation level). |
| | The policy effects are stable over time. |
| | Taxation and NW labelling are independent in influencing intake (consumption). |
| | Multiplicative risk effects. |

**Appendix Table 5.** Estimated health and economic impacts of an 8% tax rate on HFSS food without accounting for reductions in fruits and vegetables

| | **Taxation (an 8% tax rate)** |
|---|---|
| **Health impacts** | |
| Obesity | |
| - Cases | 430 000 (330 000, 530 000) |
| - Case-years | 14 000 000 (11 000 000, 17 000 000) |
| - Percentage point reduction in obesity | 2.19 (1.69, 2.73) |
| Type 2 diabetes mellitus | |
| - Cases | 100 000 (67 000, 140 000) |
| - Case-years | 500 000 (250 000, 780 000) |
| Cardiovascular diseases | |
| - Cases | 88 000 (59 000, 140 000) |
| - Case-years | 430 000 (270 000, 680 000) |
| Multiple long-term conditions | |
| - Cases | 53 000 (37 000, 75 000) |
| - Case-years | 300 000 (200 000, 430 000) |
| All-cause deaths | 24 000 (15 000, 37 000) |
| QALYs gained | 880 000 (680 000, 1 100 000) |
| **Economic impacts (in £ billion, except for BCR, ICER)** | |
| Healthcare perspective | |
| - Cost savings | 21 (17, 27) |
| - NMB (NICE's lower value) | 39 (30, 49) |
| - NMB (NICE's upper value) | 48 (37, 59) |
| - NMB (UK Government's value) | 76 (62, 96) |
| - BCR (NICE's lower value) | 1400 (1200, 1800) |
| - BCR (NICE's upper value) | 1800 (1400, 2200) |
| - BCR (UK Government's value) | 3300 (2600, 4100) |
| - ICER | 24 000 (23 000, 25 000) |
| Societal perspective | |
| - Cost savings | 53 (42, 67) |
| - NMB (NICE's lower value) | 71 (56, 89) |
| - NMB (NICE's upper value) | 80 (62, 110) |
| - NMB (UK Government's value) | 120 (90, 140) |
| - BCR (NICE's lower value) | 2800 (2200, 3500) |
| - BCR (NICE's upper value) | 3200 (2500, 4000) |
| - BCR (UK Government's value) | 4600 (3600, 5700) |
| - ICER | 61 000 (58 000, 63 000) |

*BCR = Benefit-cost ratio; ICER = Incremental cost-effectiveness ratio; NICE = National Institute for Health and Care Excellence (NICE's lower value = £20,000, NICE's upper value = £30,000, UK Government's value = £70,000); NMB = Net Monetary Benefit; QALYs = Quality-adjusted life years*
*Median and 95% uncertainty intervals (UIs) are presented, unless otherwise specified.*
*ICER is presented as a cost saving per QALY; a positive value indicates the modelled policy improves health and saves money (dominant).*

**Notes:**
Absolute equity slope index of cumulative QALYs gained : 41 000 (27 000, 59 000)
Relative equity slope index of cumulative QALYs gained : 0.05 (0.03, 0.07)

**Appendix Table 6.** Estimated health and economic impacts of an 8% tax rate on HFSS based on assumed differences in taxation effects by income levels

| | **Taxation (an 8% tax rate)** |
|---|---|
| **Health impacts** | |
| Obesity | |
| - Cases | 310 000 (230 000, 370 000) |
| - Case-years | 11 000 000 (9 600 000, 12 000 000) |
| - Percentage point reduction in obesity | 1.68 (1.53, 1.90) |
| Type 2 diabetes mellitus | |
| - Cases | 81 000 (53 000, 110 000) |
| - Case-years | 390 000 (170 000, 620 00) |
| Cardiovascular diseases | |
| - Cases | 66 000 (43 000, 92 000) |
| - Case-years | 300 000 (190 000, 450 000) |
| Multiple long-term conditions | |
| - Cases | 49 000 (36 000, 64 000) |
| - Case-years | 210 000 (140 000, 320 000) |
| All-cause deaths | 21 000 (14 000, 31 000) |
| QALYs gained | 680 000 (600 000, 750 000) |
| **Economic impacts (in £ billion, except for BCR, ICER)** | |
| Healthcare perspective | |
| - Cost savings | 16 (14, 19) |
| - NMB (NICE's lower value) | 30 (26, 34) |
| - NMB (NICE's upper value) | 37 (32, 41) |
| - NMB (UK Government's value) | 77 (67, 87) |
| - BCR (NICE's lower value) | 1200 (1100, 1300) |
| - BCR (NICE's upper value) | 1500 (1300, 1600) |
| - BCR (UK Government's value) | 2500 (2300, 2800) |
| - ICER | 24 000 (23 000, 25 000) |
| Societal perspective | |
| - Cost savings | 40 (36, 46) |
| - NMB (NICE's lower value) | 54 (48, 61) |
| - NMB (NICE's upper value) | 61 (54, 68) |
| - NMB (UK Government's value) | 88 (77, 98) |
| - BCR (NICE's lower value) | 2200 (1900, 2400) |
| - BCR (NICE's upper value) | 2400 (2100, 2700) |
| - BCR (UK Government's value) | 3500 (3100, 3900) |
| - ICER | 60 000 (58 000, 62 000) |

*BCR = Benefit-cost ratio; ICER = Incremental cost-effectiveness ratio; NICE = National Institute for Health and Care Excellence (NICE's lower value = £20,000, NICE's upper value = £30,000, UK Government's value = £70,000); NMB = Net Monetary Benefit; QALYs = Quality-adjusted life years*
*Median and 95% uncertainty intervals (UIs) are presented, unless otherwise specified.*
*ICER is presented as a cost saving per QALY; a positive value indicates the modelled policy improves health and saves money (dominant).*

**Notes:**
Absolute equity slope index of cumulative QALYs gained : 130 000 (110 000, 150 000)
Relative equity slope index of cumulative QALYs gained : 0.19 (0.17, 0.21)

**Appendix Table 7.** Estimated health and economic impacts of NW labelling on HFSS based on reductions in total energy and salt intake (as opposed to reduction per 100 grams of food)

| | **NW labelling (consumer response)** |
|---|---|
| **Health impacts** | |
| Obesity | |
| - Cases | 500 000 (330 000, 700 000) |
| - Case-years | 18 000 000 (10 000 000, 29 000 000) |
| - Percentage point reduction in obesity | 2.86 (1.68, 4.72) |
| Type 2 diabetes mellitus | |
| - Cases | 130 000 (77 000, 210 000) |
| - Case-years | 640 000 (290 000, 1 300 000) |
| Cardiovascular diseases | |
| - Cases | 110 000 (57 000, 200 000) |
| - Case-years | 550 000 (260 000, 1 000 000) |
| Multiple long-term conditions | |
| - Cases | 84 000 (47 000, 140 000) |
| - Case-years | 400 000 (210 000, 660 000) |
| All-cause deaths | 31 000 (14 000, 52 000) |
| QALYs gained | 1 100 000 (650 000, 1 800 000) |
| **Economic impacts (in £ billion, except for BCR, ICER)** | |
| Healthcare perspective | |
| - Cost savings | 28 (16, 46) |
| - NMB (NICE's lower value) | 51 (29, 83) |
| - NMB (NICE's upper value) | 62 (35, 110) |
| - NMB (UK Government's value) | 110 (61, 180) |
| - BCR (NICE's lower value) | 290 (170, 480) |
| - BCR (NICE's upper value) | 360 (200, 580) |
| - BCR (UK Government's value) | 620 (350, 1000) |
| - ICER | 24 000 (23 000, 25 000) |
| Societal perspective | |
| - Cost savings | 70 (40, 110) |
| - NMB (NICE's lower value) | 92 (53, 150) |
| - NMB (NICE's upper value) | 110 (59, 170) |
| - NMB (UK Government's value) | 150 (85, 240) |
| - BCR (NICE's lower value) | 530 (300, 870) |
| - BCR (NICE's upper value) | 600 (340, 980) |
| - BCR (UK Government's value) | 860 (490, 1400) |
| - ICER | 61 000 (59 000, 63 000) |

*BCR = Benefit-cost ratio; ICER = Incremental cost-effectiveness ratio; NICE = National Institute for Health and Care Excellence (NICE's lower value = £20,000, NICE's upper value = £30,000, UK Government's value = £70,000); NMB = Net Monetary Benefit; QALYs = Quality-adjusted life years*
*Median and 95% uncertainty intervals (UIs) are presented, unless otherwise specified.*
*ICER is presented as a cost saving per QALY; a positive value indicates the modelled policy improves health and saves money (dominant).*

**Notes:**
Absolute equity slope index of cumulative QALYs gained : 55 000 (29 000, 110 000)
Relative equity slope index of cumulative QALYs gained : 0.05 (0.03, 0.06)

**Appendix Table 8.** Absolute equity slope index and relative equity slope index of cumulative QALYs gained of an 8% tax rate, NW labelling, and both policies combined on HFSS food based on consumer response (2026 – 2040)

| | Taxation | NW labelling | Taxation and NW labelling |
|---|---|---|---|
| **QALYs gained** | | | |
| - Absolute index | 41 000 (27 000, 59 000) | 2400 (-4900, 11 000) | 46 000 (28 000, 64 000) |
| - Relative index | 0.05 (0.03, 0.06) | 0.01 (-0.02, 0.04) | 0.04 (0.02, 0.06) |

*QALYs = Quality-adjusted life years*
*The absolute equity slope index represents the policy impact on absolute inequality (i.e., QALYs gained in the most deprived areas compared to the least deprived areas).*
*The relative equity slope index represents the policy impact on absolute inequality (i.e., positive values indicate the policy tackles relative inequality).*
*Median and 95% uncertainty intervals (UIs) are presented, unless otherwise specified.*

**Appendix Table 9.** Estimated health and economic impacts of an 8% tax rate, NW labelling, and both policies combined on HFSS food based on consumer response and reformulation (2026 – 2040)

| | **Taxation** | **NW labelling** | **Taxation and NW labelling** |
|---|---|---|---|
| **Health impacts** | | | |
| Obesity | | | |
| - Cases | 430 000 (340 000, 520 000) | 130 000 (47 000, 190 000) | 440 000 (360 000, 530 000) |
| - Case-years | 13 000 000 (11 000 000, 16 000 000) | 3 400 000 (1 500 000, 5 400 000) | 15 000 000 (13 000 000, 18 000 000) |
| - Percentage point reduction in obesity | 2.16 (1.82, 2.53) | 0.48 (0.20, 0.74) | 2.43 (2.06, 2.82) |
| Type 2 diabetes mellitus | | | |
| - Cases | 99 000 (64 000, 130 000) | 26 000 (11 000, 45 000) | 120 000 (74 000, 150 000) |
| - Case-years | 480 000 (230 000, 720 000) | 130 000 (48 000, 260 000) | 570 000 (260 000, 830 000) |
| Cardiovascular diseases | | | |
| - Cases | 84 000 (59 000, 120 000) | 24 000 (10 000, 41 000) | 94 000 (64 000, 140 000) |
| - Case-years | 410 000 (250 000, 620 000) | 120 000 (44 000, 210 000) | 490 000 (300 000, 680 000) |
| Multiple long-term conditions | | | |
| - Cases | 65 000 (46 000, 84 000) | 16 000 (7500, 26 000) | 75 000 (55 000, 100 000) |
| - Case-years | 290 000 (210 000, 400 000) | 74 000 (34 000, 130 000) | 330 000 (230 000, 450 000) |
| All-cause deaths | 23 000 (16 000, 36 000) | 6100 (2200, 13 000) | 27 000 (18 000, 40 000) |
| QALYs gained | 860 000 (730 000, 1000 000) | 230 000 (97 000, 360 000) | 990 000 (830 000, 1 200 000) |
| **Economic impacts (in £ billion, except for BCR, ICER)** | | | |
| Healthcare perspective | | | |
| - Cost savings | 21 (18, 24) | 5.4 (2.4, 8.7) | 24 (20, 28) |
| - NMB (NICE's lower value) | 36 (31, 43) | 9 (3.5, 15) | 42 (35, 49) |
| - NMB (NICE's upper value) | 45 (38, 53) | 11 (4.4, 19) | 52 (44, 61) |
| - NMB (UK Government's value) | 79 (67, 93) | 20 (8.3, 33) | 92 (70, 110) |
| - BCR (NICE's lower value) | 25 (21, 29) | 11 (4.9, 18) | 27 (22, 31) |
| - BCR (NICE's upper value) | 31 (26, 36) | 14 (6, 22) | 33 (27, 38) |
| - BCR (UK Government's value) | 53 (45, 62) | 24 (10, 38) | 57 (48, 66) |
| - ICER | 22 000 (22 000, 23 000) | 20 000 (16 000, 22 000) | 23 000 (22 000, 24 000) |
| Societal perspective | | | |
| - Cost savings | 52 (44, 61) | 13 (6, 22) | 60 (51, 70) |

| | | | |
|---|---|---|---|
| - NMB (NICE's lower value) | 67 (57, 80) | 17 (7, 28) | 79 (66, 91) |
| - NMB (NICE's upper value) | 76 (64, 90) | 19 (8, 32) | 88 (74, 100) |
| - NMB (UK Government's value) | 110 (93, 130) | 29 (12, 46) | 130 (110, 150) |
| - BCR (NICE's lower value) | 46 (39, 54) | 20 (8.9, 33) | 49 (41, 56) |
| - BCR (NICE's upper value) | 51 (43, 60) | 23 (10, 37) | 55 (46, 63) |
| - BCR (UK Government's value) | 74 (63, 87) | 33 (14, 53) | 79 (66, 91) |
| - ICER | 59 000 (57 000, 61 000) | 57 000 (52 000, 60 000) | 59 000 (57 000, 61 000) |

*BCR = Benefit-cost ratio; ICER = Incremental cost-effectiveness ratio; NICE = National Institute for Health and Care Excellence (NICE's lower value = £20,000, NICE's upper value = £30,000, UK Government's value = £70,000); NMB = Net Monetary Benefit; QALYs = Quality-adjusted life years*

*Median and 95% uncertainty intervals (UIs) are presented, unless otherwise specified.*

*ICER is presented as a cost saving per QALY; a positive value indicates the modelled policy improves health and saves money (dominant).*

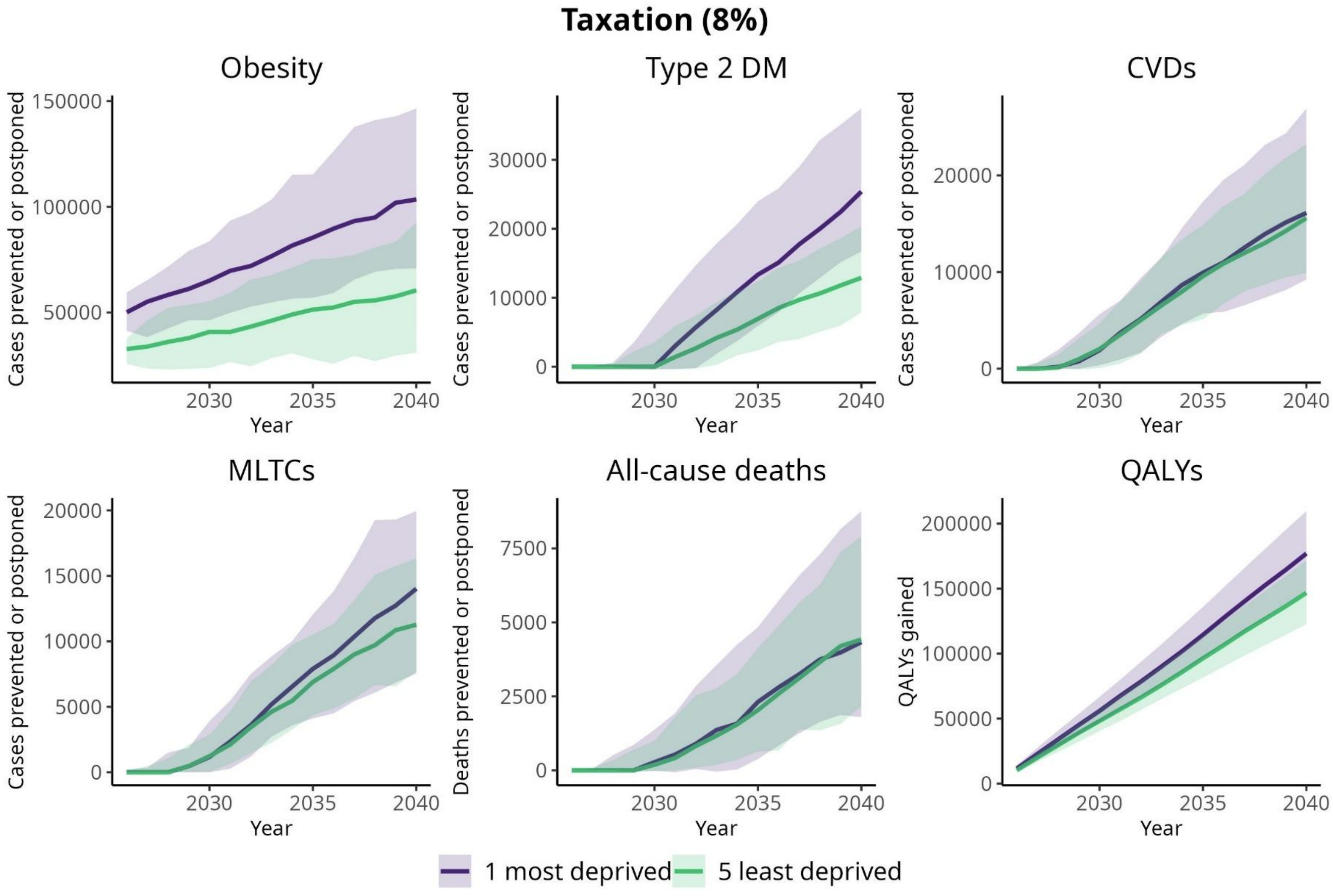

**Appendix Figure 3.** Cumulative cases or deaths prevented or postponed and QALYs gained (median; 95% UI) due to taxation (8%) on HFSS food based on consumer response and reformulation by QIMD (2026 – 2040)

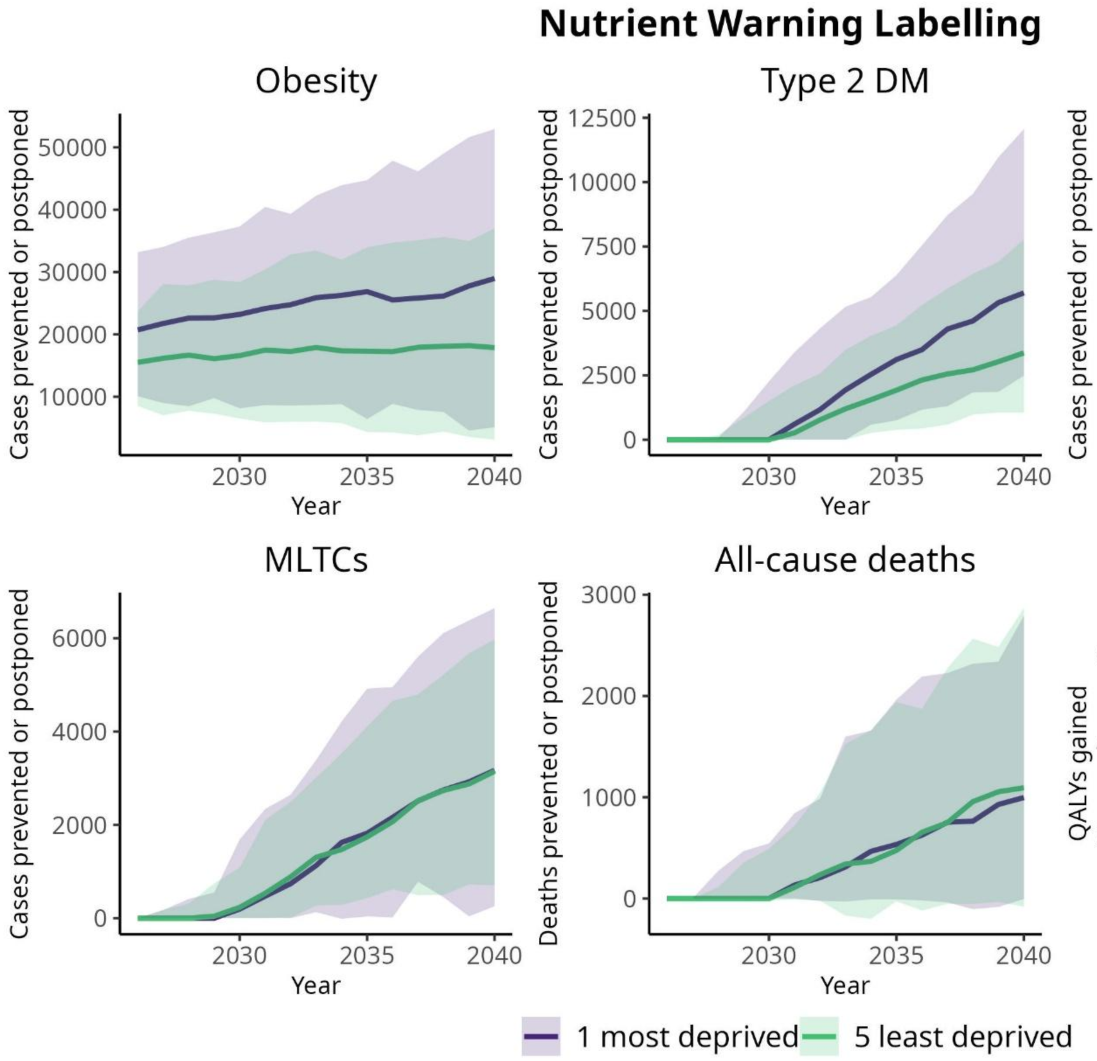

**Appendix Figure 4.** Cumulative cases or deaths prevented or postponed and QALYs gained (median; 95% UI) due to NW labelling on HFSS food based on consumer response and reformulation by QIMD (2026 – 2040)

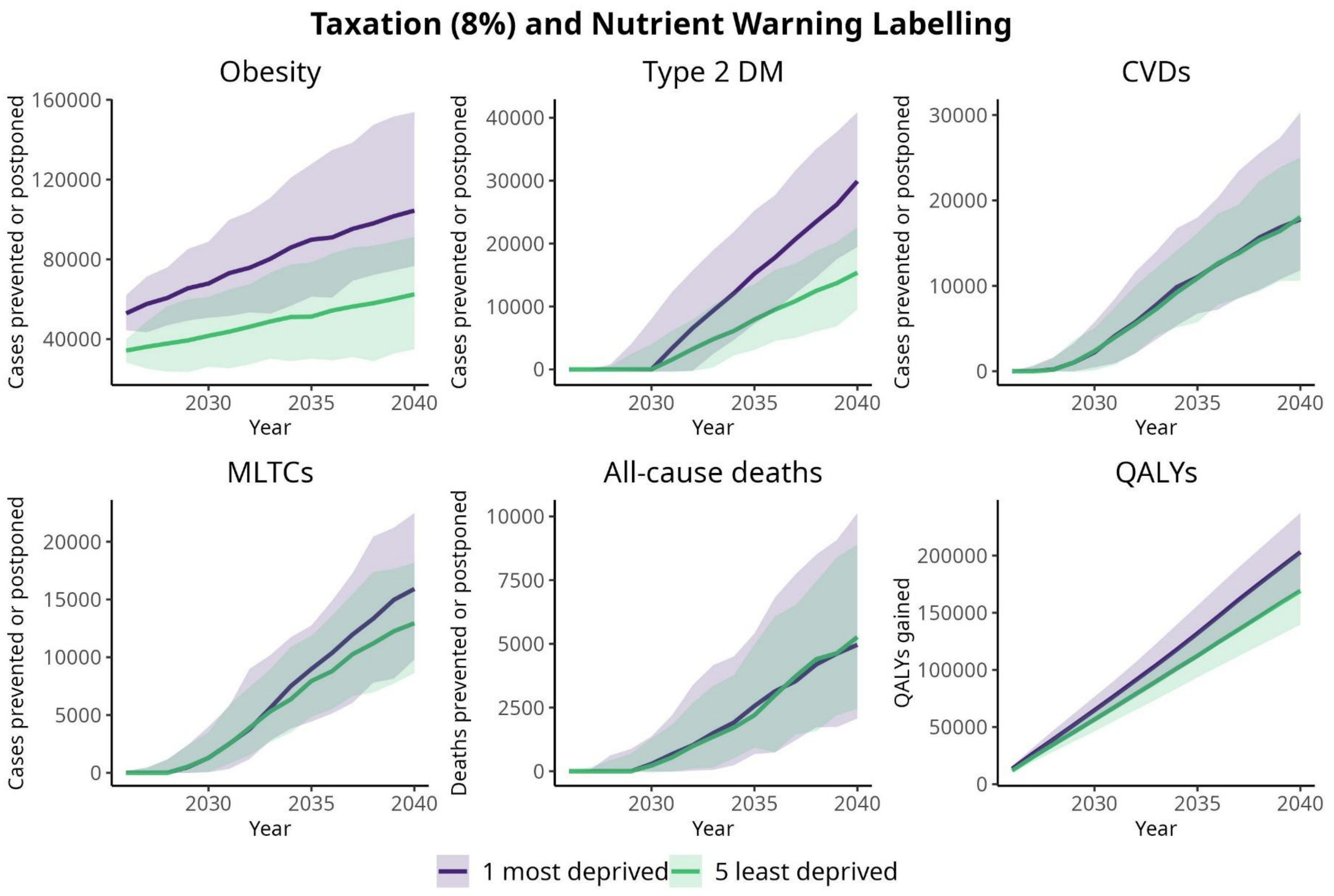

**Appendix Figure 5.** Cumulative cases or deaths prevented or postponed and QALYs gained (median; 95% UI) due to taxation (8%) and NW labelling on HFSS food based on consumer response and reformulation by QIMD (2026 – 2040)

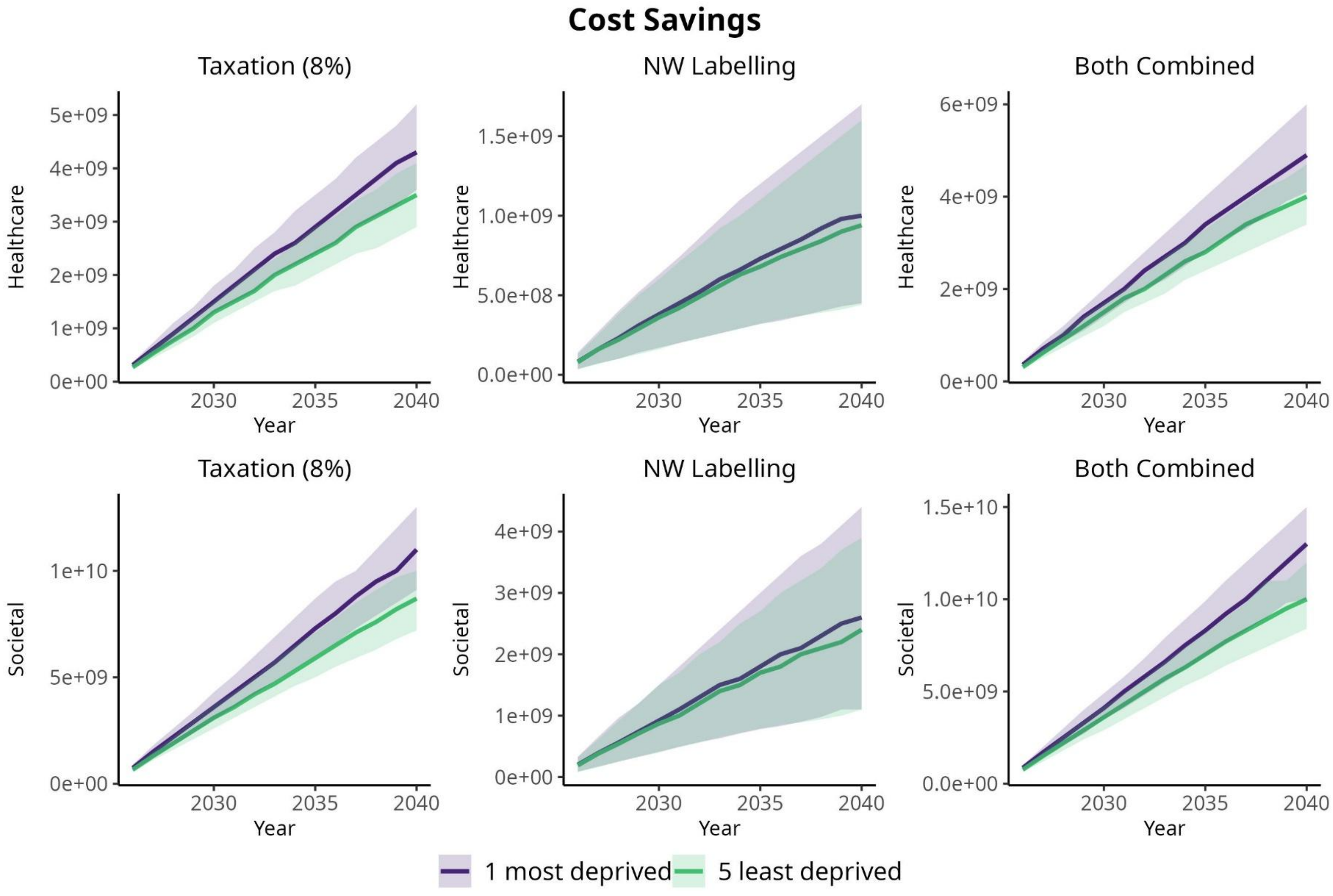

**Appendix Figure 6.** Cumulative cost savings (median; 95% UI) due to policy options based on consumer response and reformulation by QIMD (2026 – 2040)

**Appendix Table 10.** Absolute equity slope index and relative equity slope index of cumulative QALYs gained of an 8% tax rate, NW labelling, and both policies combined on HFSS food based on consumer response and reformulation (2026 – 2040)

| | Taxation | NW labelling | Taxation and NW labelling |
|---|---|---|---|
| **QALYs gained** | | | |
| - Absolute index | 41 000 (25 000, 57 000) | 3100 (-3000, 11 000) | 43 000 (26 000, 58 000) |
| - Relative index | 0.05 (0.03, 0.07) | 0.01 (-0.01, 0.05) | 0.04 (0.03, 0.06) |

*QALYs = Quality-adjusted life years*
*The absolute equity slope index represents the policy impact on absolute inequality (i.e., QALYs gained in the most deprived areas compared to the least deprived areas).*
*The relative equity slope index represents the policy impact on absolute inequality (i.e., positive values indicate the policy tackles relative inequality).*
*Median and 95% uncertainty intervals (UIs) are presented, unless otherwise specified.*

**Appendix Table 11.** Estimated health and economic impacts of an 8% tax rate, NW labelling, and both policies combined on HFSS food based on consumer response with compensation on energy intake (11%) (2026 – 2040)

| | Taxation | NW labelling | Taxation and NW labelling |
|---|---|---|---|
| **Health impacts** | | | |
| Obesity | | | |
| - Cases | 410 000 (310 000, 490 000) | 110 000 (43 000, 190 000) | 430 000 (330 000, 520 000) |
| - Case-years | 12 000 000 (9 500 000, 15 000 000) | 3 000 000 (1 300 000, 5 400 000) | 15 000 000 (12 000 000, 19 000 000) |
| - Percentage point reduction in obesity | 1.97 (1.50, 2.42) | 0.42 (0.17, 0.75) | 2.36 (1.86, 2.93) |
| Type 2 diabetes mellitus | | | |
| - Cases | 89 000 (59 000, 120 000) | 23 000 (11 000, 46 000) | 110 000 (73 000, 150 000) |
| - Case-years | 450 000 (220 000, 680 000) | 120 000 (48 000, 250 000) | 560 000 (260 000, 860 000) |
| Cardiovascular diseases | | | |
| - Cases | 73 000 (51 000, 120 000) | 21 000 (9000, 37 000) | 94 000 (62 000, 130 000) |
| - Case-years | 370 000 (230 000, 610 000) | 110 000 (39 000, 200 000) | 470 000 (280 000, 690 000) |
| Multiple long-term conditions | | | |
| - Cases | 57 000 (43 000, 84 000) | 15 000 (6600, 26 000) | 72 000 (52 000, 98 000) |
| - Case-years | 260 000 (170 000, 390 000) | 68 000 (29 000, 130 000) | 330 000 (220 000, 460 000) |
| All-cause deaths | 20 000 (12 000, 33 000) | 5500 (2000, 12 000) | 25 000 (17 000, 41 000) |
| QALYs gained | 780 000 (610 000, 970 000) | 200 000 (87 000, 350 000) | 980 000 (770 000, 1 200 000) |
| **Economic impacts (in £ billion, except for BCR, ICER)** | | | |
| Healthcare perspective | | | |
| - Cost savings | 19 (15, 24) | 4.9 (2.2, 8.7) | 24 (18, 29) |
| - NMB (NICE's lower value) | 35 (27, 43) | 8.8 (3.8, 16) | 43 (34, 53) |
| - NMB (NICE's upper value) | 42 (31, 51) | 11 (4.6, 19) | 53 (41, 65) |
| - NMB (UK Government's value) | 74 (57, 91) | 19 (8.1, 33) | 92 (72, 110) |
| - BCR (NICE's lower value) | 1400 (1100, 1700) | 52 (23, 90) | 220 (170, 280) |
| - BCR (NICE's upper value) | 1700 (1300, 2100) | 63 (28, 110) | 270 (210, 340) |
| - BCR (UK Government's value) | 3000 (2300, 3600) | 110 (48, 190) | 480 (370, 590) |
| - ICER | 24 000 (23 000, 25 000) | 24 000 (22 000, 25 000) | 24 000 (23 000, 25 000) |
| Societal perspective | | | |
| - Cost savings | 47 (37, 59) | 12 (5.5, 22) | 59 (46, 73) |

| | | | |
|---|---|---|---|
| - NMB (NICE's lower value) | 63 (49, 79) | 16 (7.1, 28) | 79 (61, 97) |
| - NMB (NICE's upper value) | 71 (55, 88) | 18 (7.9, 32) | 89 (69, 110) |
| - NMB (UK Government's value) | 110 (79, 130) | 26 (11, 46) | 130 (100, 160) |
| - BCR (NICE's lower value) | 2500 (2000, 3100) | 94 (42, 160) | 410 (320, 500) |
| - BCR (NICE's upper value) | 2800 (2200, 3500) | 110 (47, 180) | 460 (360, 560) |
| - BCR (UK Government's value) | 4100 (3200, 5100) | 150 (67, 270) | 660 (520, 810) |
| - ICER | 61 000 (59 000, 62 000) | 60 000 (57 000, 62 000) | 61 000 (59 000, 62 000) |

*BCR = Benefit-cost ratio; ICER = Incremental cost-effectiveness ratio; NICE = National Institute for Health and Care Excellence (NICE's lower value = £20,000, NICE's upper value = £30,000, UK Government's value = £70,000); NMB = Net Monetary Benefit; QALYs = Quality-adjusted life years*

*Median and 95% uncertainty intervals (UIs) are presented, unless otherwise specified.*

*ICER is presented as a cost saving per QALY; a positive value indicates the modelled policy improves health and saves money (dominant).*

**Appendix Table 12.** Absolute equity slope index and relative equity slope index of cumulative QALYs gained of an 8% tax rate, NW labelling, and both policies combined on HFSS food based on consumer response with compensation on energy intake (11%) (2026 – 2040)

| | **Taxation** | **NW labelling** | **Taxation and NW labelling** |
|---|---|---|---|
| **QALYs gained** | | | |
| - Absolute index | 38 000 (23 000, 55 000) | 2000 (-4700, 9800) | 41 000 (25 000, 57 000) |
| - Relative index | 0.05 (0.03, 0.07) | 0.01 (-0.02, 0.04) | 0.04 (0.02, 0.06) |

*QALYs = Quality-adjusted life years*
*The absolute equity slope index represents the policy impact on absolute inequality (i.e., QALYs gained in the most deprived areas compared to the least deprived areas).*
*The relative equity slope index represents the policy impact on absolute inequality (i.e., positive values indicate the policy tackles relative inequality).*
*Median and 95% uncertainty intervals (UIs) are presented, unless otherwise specified.*
